# Gut microbiome shifts in adolescents after sleeve gastrectomy with increased oral-associated taxa and pro-inflammatory potential

**DOI:** 10.1101/2024.09.16.24313738

**Authors:** Cynthia O Akagbosu, Kathryn E McCauley, Sivaranjani Namasivayam, Hector N Romero-Soto, Wade O’Brien, Mickayla Bacorn, Eric Bohrnsen, Benjamin Schwarz, Shreni Mistry, Andrew S Burns, P. Juliana Perez-Chaparro, Qing Chen, Phoebe LaPoint, Anal Patel, Lauren E Krausfeldt, Poorani Subramanian, Brian A Sellers, Foo Cheung, Richard Apps, Iyadh Douagi, Shira Levy, Evan P Nadler, Suchitra K Hourigan

**Author notes:** These authors contributed equally (Cynthia O Akagbosu, Kathryn E McCauley, Sivaranjani Namasivayam). These authors contributed equally (Evan P Nadler, Suchitra K Hourigan). **Corresponding Author:** Suchitra K Hourigan. Clinical Microbiome Unit. National Institute of Allergy and Infectious Diseases, National Institutes of Health, Bethesda, Maryland, United States.

## Abstract

**Background:** Bariatric surgery is highly effective in achieving weight loss in children and adolescents with severe obesity, however the underlying mechanisms are incompletely understood, and gut microbiome changes are unknown.

**Objectives:** 1) To comprehensively examine gut microbiome and metabolome changes after laparoscopic vertical sleeve gastrectomy (VSG) in adolescents and 2) to assess whether the microbiome/metabolome changes observed with VSG influence phenotype using germ-free murine models.

**Design:** 1) A longitudinal observational study in adolescents undergoing VSG with serial stool samples undergoing shotgun metagenomic microbiome sequencing and metabolomics (polar metabolites, bile acids and short chain fatty acids) and 2) a human-to-mouse fecal transplant study.

**Results:** We show adolescents exhibit significant gut microbiome and metabolome shifts several months after VSG, with increased alpha diversity and notably with enrichment of oral-associated taxa. To assess causality of the microbiome/metabolome changes in phenotype, pre-VSG and post-VSG stool was transplanted into germ-free mice. Post-VSG stool was not associated with any beneficial outcomes such as adiposity reduction compared pre-VSG stool. However, post-VSG stool exhibited an inflammatory phenotype with increased intestinal Th17 and decreased regulatory T cells. Concomitantly, we found elevated fecal calprotectin and an enrichment of proinflammatory pathways in a subset of adolescents post-VSG.

**Conclusion:** We show that in some adolescents, microbiome changes post-VSG may have inflammatory potential, which may be of importance considering the increased incidence of inflammatory bowel disease post-VSG.

**What is already known on this topic:** Bariatric surgery is highly effective in achieving weight loss in children and adolescents with severe obesity, however the underlying mechanisms are incompletely understood, and gut microbiome changes are unknown.

**What this study adds:** Significant gut microbiome and metabolome shifts were found several months after vertical sleeve gastrectomy in adolescents, notably with enrichment of oral-associated taxa. Using human to germ-free mice fecal transplant studies, the post-surgery changes in the gut microbiome/metabolome were shown to have inflammatory potential. Furthermore, raised fecal calprotectin and inflammatory systemic pathways were seen in a subset of adolescents post-surgery.

**How this study might affect research, practice or policy:** These findings may be of importance given the growing recognition of an increased incidence of inflammatory bowel disease after bariatric surgery and warrants further investigation.

## INTRODUCTION

The epidemic of childhood obesity continues unabated, with 19.3% of children and adolescents having obesity and 6.1% having severe obesity in the USA^1^. Multiple comorbidities are associated with childhood obesity including type 2 diabetes mellitus (T2DM), metabolic dysfunction-associated steatotic liver disease (MASLD), dyslipidemia, and continuation of obesity into adulthood^2^ ^3^. If weight loss can be achieved prior to entering adulthood, the risk of these conditions is mitigated. Therefore, childhood obesity is a key target for intervention^4^.

Bariatric surgery achieves significant weight loss and reduces or resolves associated comorbidities in children with severe obesity^5^. Clinical guidelines recommend bariatric surgery as a potential intervention for children and adolescents with class II obesity and comorbidities or class III obesity with or without comorbidities^6^ ^7^. Laparoscopic vertical sleeve gastrectomy (VSG), with removal of 80-90% of the greater curvature of the stomach, is the most common pediatric bariatric procedure and the only type performed on those <13 years of age^8^.

Although VSG in children and adolescents is highly effective, the biological mechanisms underlying the weight loss and metabolic improvement are not fully elucidated. Restriction of food intake by reduction of stomach capacity plays an important role but the degree of enhanced metabolism post-VSG cannot be explained by caloric restriction alone. VSG also leads to weight loss through altered neurohormonal feedback mechanisms including increases in glucagon-like peptide-1 (GLP-1) and peptide YY, which reduces appetite^9^.

There has been growing interest in the role of the gut microbiome in the mechanisms behind VSG. Many studies have shown a difference in the gut microbiome between individuals with and without obesity ^10^. Moreover, murine models with fecal transplant (FT) from humans with obesity to germ-free mice have shown transfer of the obesity phenotype indicating a causal role for the gut microbiome^11^. Studies examining gut microbiome changes after bariatric surgery in adults suggest an increase in microbiota diversity and decrease in Firmicutes/Bacteroidetes ratio, although specific changes differ between surgery type and amongst studies^12^ ^13^.

To our knowledge, the role of the gut microbiome in adolescents undergoing bariatric surgery has not yet been examined^14^. This is uniquely important to study as the microbiome of children and adolescents differs from adults^15-17^. The developing microbiome in childhood also plays a clear role in establishing metabolic and inflammatory pathways that could impact energy regulation in obesity^18^ ^19^. Additionally, there is mounting evidence suggesting that the age of onset of obesity significantly impacts overall cardiometabolic risk, with childhood obesity possibly representing a more virulent form of the disease as both MASLD and T2DM progress more rapidly in children than adults^20-22^. Thus, studying microbiome changes in children and adolescents may be of particular importance. Furthermore, many of the studies in adults undergoing bariatric surgery focused on Roux-en-Y gastric bypass, not VSG, used 16S rRNA gene microbiome sequencing, and did not examine microbiome function and metabolites. Also, there are limited studies examining whether the microbiota changes found with bariatric surgery have a causal role in the beneficial phenotype changes seen post-surgery.

Therefore, we aimed to 1) deeply examine gut microbiome structure and function changes with VSG in adolescents using shotgun metagenomic sequencing and wide metabolomic analysis and 2) assess if the microbiome/metabolome differences seen have a causal role in the phenotypic changes observed with VSG by performing FT of stool from adolescents pre-VSG and post-VSG into germ-free mice.

## RESULTS

### Participants and clinical data

Twelve participants provided paired stool samples within the 8 weeks (mean 2 weeks) prior to VSG (pre-VSG) and follow-up stool samples 3-7 months (mean 5 months) after VSG (post-VSG). The mean age at VSG was 15 years (range 10-18 years), 8/12 participants were female and 9/12 Black or African American (Table 1). Notably, 2 participants were identical twins.

**Table 1:** Subject demographics, clinical data, and changes with laparoscopic vertical sleeve gastrectomy (VSG). Significant p-values in bold.

|  | **Pre-VSG**<br>**(n=12)** | **Post-VSG**<br>**(n=12)** | **P-value** |
| --- | --- | --- | --- |
| **Demographics** |  |  |  |
| Age in years, mean (range) | 15 (10-18) |  |  |
| Female Sex | 8 (67%) |  |  |
| Race: |  |  |  |
| White or Caucasian | 2 (17%) |  |  |
| Black or African American | 9 (75%) |  |  |
| Other | 1 (8%) |  |  |
| Hispanic or Latino Ethnicity | 1 (8%) |  |  |
| **Clinical** |  |  |  |
| Body mass index (BMI), kg/m<sup>2</sup> | 48.7 | 39.9 | **<0.0001** |
| Weight in kg, mean (range) | 144 (94-195) | 120 (67–173) | **<0.0001** |
| Diabetes or Prediabetes | 8 (3 with diabetes, 5 with prediabetes) | 0 | **0.0078** |
| Dyslipidemia | 4 | 1 | 0.25 |
| Hypertension | 4 | 3 | 1 |
| HbA1c % (range) | 5.6 (5.3-5.9) | 5.2 (4.9-5.5) | **0.0029** |
| Low-density lipoprotein cholesterol mg/dL (range) | 100.9 (64-157) | 90.4 (66-126) | 0.947 |
| High-density lipoprotein cholesterol mg/dL (range) | 41.3 (29-58) | 49 (42-61) | **0.0317** |
| Triglycerides mg/dL (range) | 111.5 (56-195) | 81 (39-114) | 0.271 |
| Alanine aminotransferase U/L (range) | 28.2 (9-59) | 15.7 (6-28) | **0.0025** |

At VSG, subjects had a mean body mass index (BMI) of 48.7kg/m^2^ which decreased to 39.9 kg/m^2^ (p<0.0001) post-VSG (Table 1). Total body weight loss (TBWL) averaged 17.8% (range 5.9% - 32.9%). 8/12 participants had T2DM or prediabetes pre-VSG with a reduction to 0/12 post-VSG (p=0.0078). 7/12 participants had elevated alanine aminotransferase (ALT) pre-VSG, indicating a high likelihood of MASLD; only 1/12 participants had a liver biopsy which showed metabolic dysfunction-associated steatohepatitis. Overall, mean ALT decreased from 28.2 U/L to 15.7U/L (p=0.0025) with VSG. 4/12 participants had dyslipidemia with elevated low-density lipoprotein (LDL) pre-VSG compared to 1/12 post-VSG, with an increase in high-density lipoprotein (HDL) cholesterol post-VSG from an average of 41.3 mg/dL to 49.0 mg/dL (p=0.0317).

### Stool microbiome changes with VSG

#### i. Alpha and beta diversity

Pre-VSG and post-VSG stool samples underwent shotgun metagenomic sequencing. Bacterial diversity increased post-VSG (Shannon p=0.047, Inv Simpson p=0.04, Evenness (p=0.042) Fig.1A). Significant changes in microbiome composition (beta diversity) were seen using the Canberra distance, which places more weight on lower abundance species (p=0.015, Fig.1B) but not Bray Curtis distance. Pre-VSG, alpha diversity was significantly lower in those with diabetes compared with pre-diabetes and no diabetes (SuppFig.1A) and microbiome composition also differed (SuppFig.1B). Post-VSG, there were no differences in alpha or beta diversity between those who previously had diabetes and the other subjects. There were no significant differences between changes in alpha and beta diversity and other clinical parameters.

**Fig.1.**
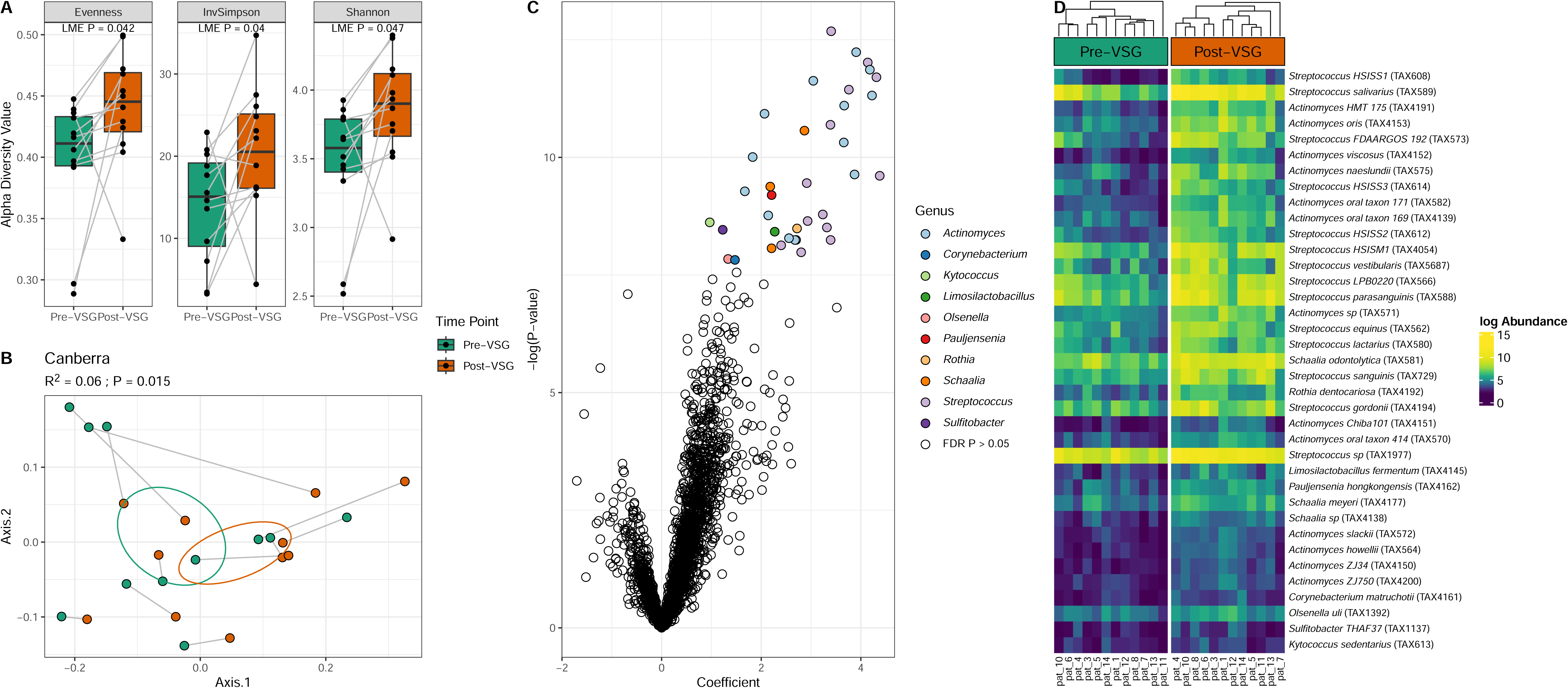
Microbial taxa exhibit significant changes after VSG. (A) Subjects exhibited increases in several alpha diversity metrics post-VSG (LME P-value < 0.05). (B) Significant changes in underlying microbiota were also observed (Canberra PERMANOVA R^2^ = 0.06, P=0.015). (C) Taxa enriched post-VSG included several members of the *Streptococcus* and *Actinomyces* genera (FDR P < 0.05). (D) All taxa at a species level that significantly increased post-VSG in order of coefficient. Abbreviations: VSG: vertical sleeve gastrectomy, FDR: False Discovery Rate, LME: Linear Mixed Effects; PERMANOVA: Permutational Analysis of Variance.

#### ii. Taxonomic shifts

There was a significant enrichment of 76 bacterial taxa post-VSG. Notably, the top 18 species enriched post-VSG were from the *Streptococcus* and *Actinomyces* genera (Fig.1C-1D. Supp Table 1). This included an enrichment of *Streptococcus salivarius, Streptococcus vestibularis, Streptococcus parasanguinis, Actinomyces oris* and *Actinomyces oral taxon*, all of which are commonly associated with the oral cavity. No individual taxa significantly correlated with clinical characteristics.

#### iii. Carbohydrate-Active Enzymes (CAZymes)

CAZymes were examined due to their role in influencing host metabolism^23^. While the overall composition of CAZymes only showed a moderate change with VSG (Canberra PERMANOVA R2 = 0.049, P=0.056, Fig.2A), five specific CAZymes exhibited significant enrichment post-VSG (Fig.2B, Supp Table 2). Many of the CAZyme enrichments post-VSG were associated with *Streptococcus species* (Fig.2C). In addition, Glycoside Hydrolase 13 (GH13)+Carbohydrate Binding Module 20 (CBM 20) significantly associated with increased TBWL (q=0.002, Supp Table 2).

**Fig.2.**
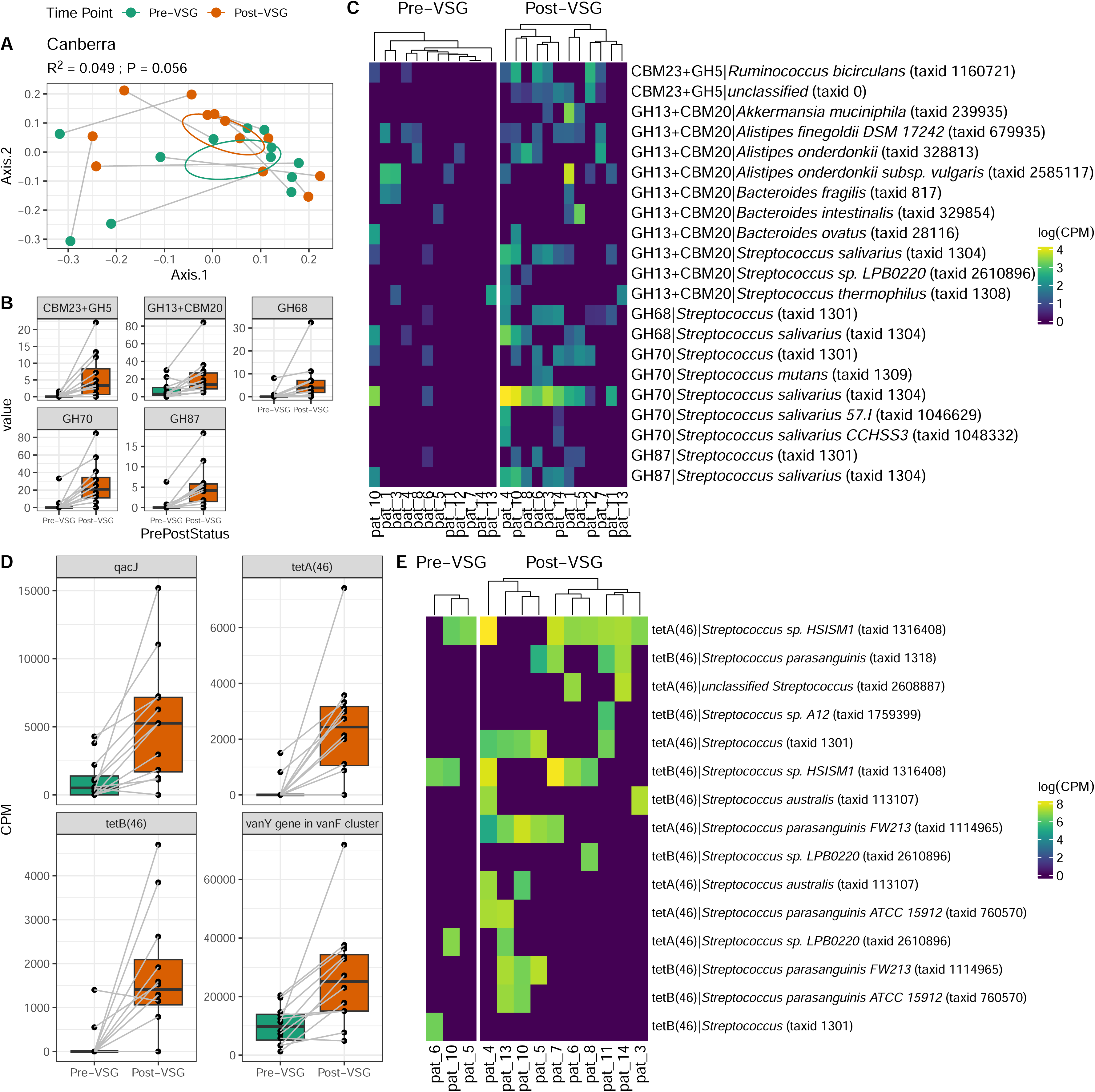
Carbohydrate-Active Enzymes (CAZymes) and Antibiotic Resistance Genes (ARGs) identified from metagenomic data exhibit increases in *Streptococcus* species after VSG. (A) Composition of CAZymes shows moderate change (Canberra PERMANOVA R2 = 0.049, P=0.056). (B) Interrogating specific CAZymes, five exhibited significant enrichments post-VSG (MaAsLin2, FDR P < 0.25). (C) Heatmap shows significant CAZyme genes from panel B, stratified by gene-based taxonomy. (D) Antibiotic Resistance Genes enriched (Pfdr < 0.1) after surgery. (E) Antibiotic resistance genes with the largest effect size, tetA(46) and tetB(46) identified primarily in Streptococcus contiguous sequences. Abbreviations: VSG: vertical sleeve gastrectomy, CBM: Carbohydrate Binding Module, CPM: Copies Per Million, GH: Glycoside Hydrolase, PERMANOVA: Permutational Analysis of Variance.

#### iv. Antibiotic resistance genes (ARGs)

ARGs were examined using the RGI CARD database. Four ARGs were enriched post-VSG: qacJ, tetA(46), tetB(46) and vanY in vanF cluster (Fig.2D). These resistance genes, especially tetAB(46), were primarily found in contigs belonging to several *Streptococcus* species enriched post-VSG (Fig.2E).

#### v. Conserved functional enrichments

There were no functional pathway level differences with VSG. Therefore, reads were aligned to Enzyme Commission (EC) gene annotations to identify more refined gene function differences. There was a moderate change in lower-abundance genes post-VSG (Canberra R2=0.05, P=0.059, SuppFig.2A), most notably with enrichment of EC 1.1.1.105, an all-trans retinol dehydrogenase gene, from the oxidoreductases class (SuppFig.2B, Supp Table 3). In addition, these enriched ECs formed three distinct modules of co-associated genes (blue, brown and turquoise) that increased significantly post-VSG (SuppFig.2C-D), driven by different bacterial genera (SuppFig.2E). ECs in the blue module were primarily from mevalonate, hemiterpene biosynthesis and heme biosynthesis pathways; those in the brown module focused on sugar acid degradation, and the turquoise module contained several tRNA synthetases (Supp Table 3).

#### vi. Bacteriophage and viral composition

When looking at the DNA virome, there were no significant differences observed with VSG in taxonomy, host taxonomy, taxonomic diversity, taxonomic composition, viral protein family diversity or composition (SuppFig.3A-F). As most DNA viruses in the gut microbiome are phages, this suggests phage-containing bacteria did not significantly change with VSG despite large overall bacterial changes.

**Fig.3.**
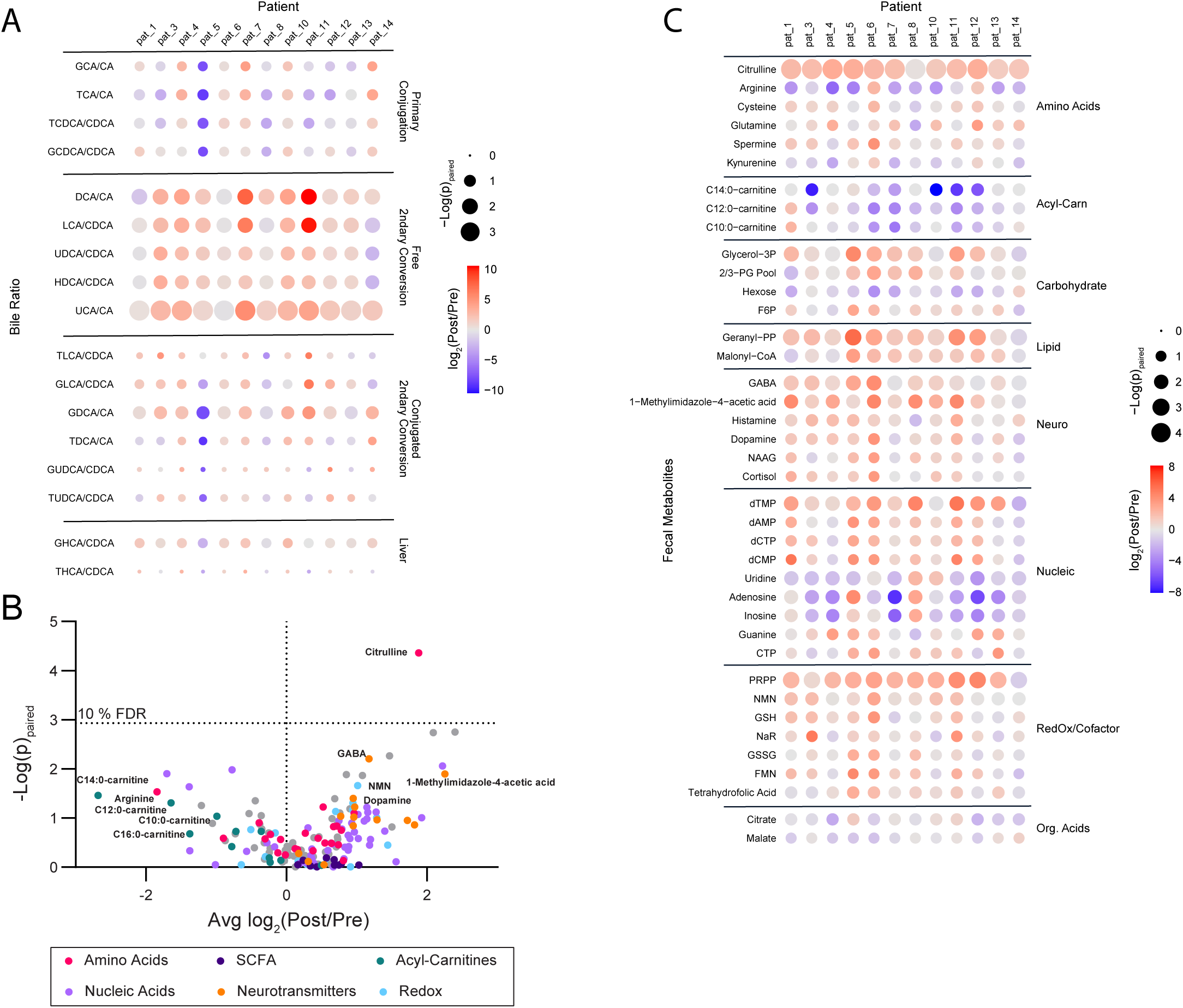
Stool metabolite changes with VSG. (A) Bile acid levels assessed by LC-MS/MS and the conversion ratios were calculated to reflect known primary to secondary bile acid, and unconjugated to conjugated bile acid substrate/product pairs. Ratios post- vs. pre-VSG were analyzed by a paired t-test and the fold change and significance of the change are reflected as the color of each node and the size of each node by row respectively in the displayed heatmap. All measured ratios are displayed. (B) Polar fecal metabolites were assessed by LC-MS/MS and total sum normalized prior to analysis. Paired fold changes of post- vs pre-VSG and significance by paired t-test are displayed with a false discovery rate of 10 % indicated at p=0.0012, calculated via a Benjamini-Hochberg correction. Pathways and molecular families of interest are indicated via color. (C) A heatmap of the paired post- vs. pre-VSG changes displayed in (B) broken out by participant to display variance and observed pathway-driven trends in metabolite levels. All displayed features have a raw p<0.1 via a paired t-test. Abbreviations: LC-MS/MS: Liquid Chromatography Mass Spectrometry, VSG: vertical sleeve gastrectomy, Acyl-Carn: acyl-carnitines, Neuro: neurotransmitters, Nucleic: nucleic acids, nucleosides, and nucleotides, Org. Acids: organic acids, SCFA: short-chain fatty acids

### Stool metabolome changes with VSG

Post-VSG stool displayed higher ratios of secondary to primary bile acids compared to pre-VSG stool (Fig.3A). Notably, this pattern was broad and included all ratios for which the corresponding precursor primary bile acid and product secondary bile acid were detected.

Amongst polar metabolites, only the elevation of citrulline post-VSG passed a false discovery rate cut-off of 10% (Fig.3B). However, several trends known to be important to gut health were observed (Fig.3C). These included a decrease in acyl-carnitines, an increase in neurotransmitters known to be directly microbially produced (GABA, dopamine, and histamine), and increases in redox cofactor metabolites. No changes in SCFAs were observed.

Polar metabolites were related to a range of demographic and clinical factors (SuppFig.4, Supp Table 4). Several amino acid metabolites (purple) positively associated with HDL, and negatively associated with hemoglobin A1C (HbA1c) pre-VSG. In addition, several metabolites correlated negatively with BMI pre-VSG, including those in Redox and Co-Enzyme metabolite (CoA, FMN, NADH), Glycolysis (G1P, Glycerol-3P), and Nucleotide (UMP, AMP) classes. Post-VSG, several metabolites associated with the degree of TBWL, including neurotransmitters such as GABA and Dopamine, which also correlated negatively with triglyceride levels. Citrulline did not correlate with any factors pre-VSG or post-VSG.

**Fig.4.**
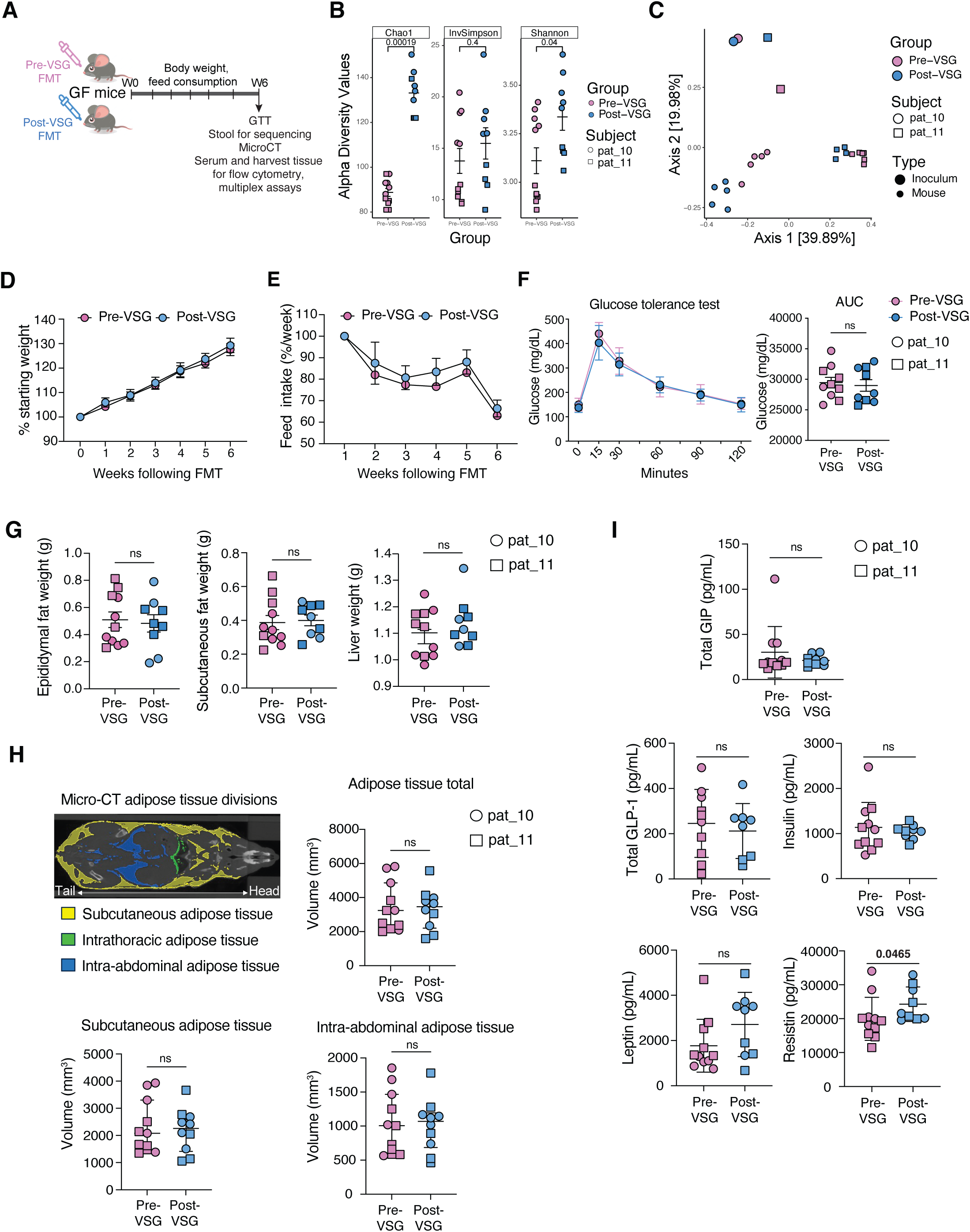
Fecal transplant with pre-VSG and post-VSG stool into germ-free mice. (A) Schematic of experimental design. Pre-VSG and post-VSG stool samples were inoculated (FT) into two groups of germ-free mice (n=4-6/group). Body weights and feed consumption were measured every week and mouse stool samples collected at 6 weeks were sequenced and analyzed. At week 6 following FT, a glucose tolerance test was performed. Following micro-CT imaging, mice were sacrificed for sample collection. The experiment was repeated using samples from two participants. (B) Alpha diversity was higher among mice receiving post-VSG stool, compared to those receiving pre-VSG stool. (C) Mice also exhibited compositional differences between pre-VSG and post-VSG stool, which was distinct from inoculum samples. (D)Weight change in mice following FMT with pre-VSG and post-VSG stool is shown as a percentage of starting weight. No significant differences were found. (E) No differences in feed consumption between the two groups were found. (F) Glucose tolerance tests showed blood glucose levels at different time points of the test (left) and the area under the curve for the entire test is plotted (right) for the two groups. The two participants are identified as shown in the key. (G) Weight measurements for epididymal and subcutaneous fat pads and liver tissues are plotted for the two groups. (H) Micro-CT results showing adipose areas examined in the mouse (top left), total adipose tissue volume (top right), subcutaneous adipose tissue volume (bottom left), and intraabdominal adipose tissue volume (bottom right). No differences were seen between groups. (I) Serum metabolic hormone measurements. Resistin was higher following FT with post-VSG stool compared with pre-VSG stool; no other differences were seen. Abbreviations: VSG: vertical sleeve gastrectomy, FT: fecal transplant.

### Stool microbiome and metabolite correlations with VSG

To reduce the dimensionality of the microbiota, taxa were agglomerated into seven co-associated networks. Each increased in abundance post-VSG except for the red network, which exhibited a trending decrease (p=0.087; SuppFig.5A-B, Supp Table 5). Changes in these networks were correlated with changes in polar metabolites which revealed that the *Bacteroides* and the *Alistipes/Akkermansia/Actinomyces* prominent group both increased in concert with several SCFAs including butyrate, isovalerate, and isobutyrate (SuppFig.5C, Supp Table 5). Conversely, the *Streptococcus* prominent group had the fewest correlations, only exhibiting positive correlations with isocitrate and cytidine and a negative correlation with urate.

**Fig.5.**
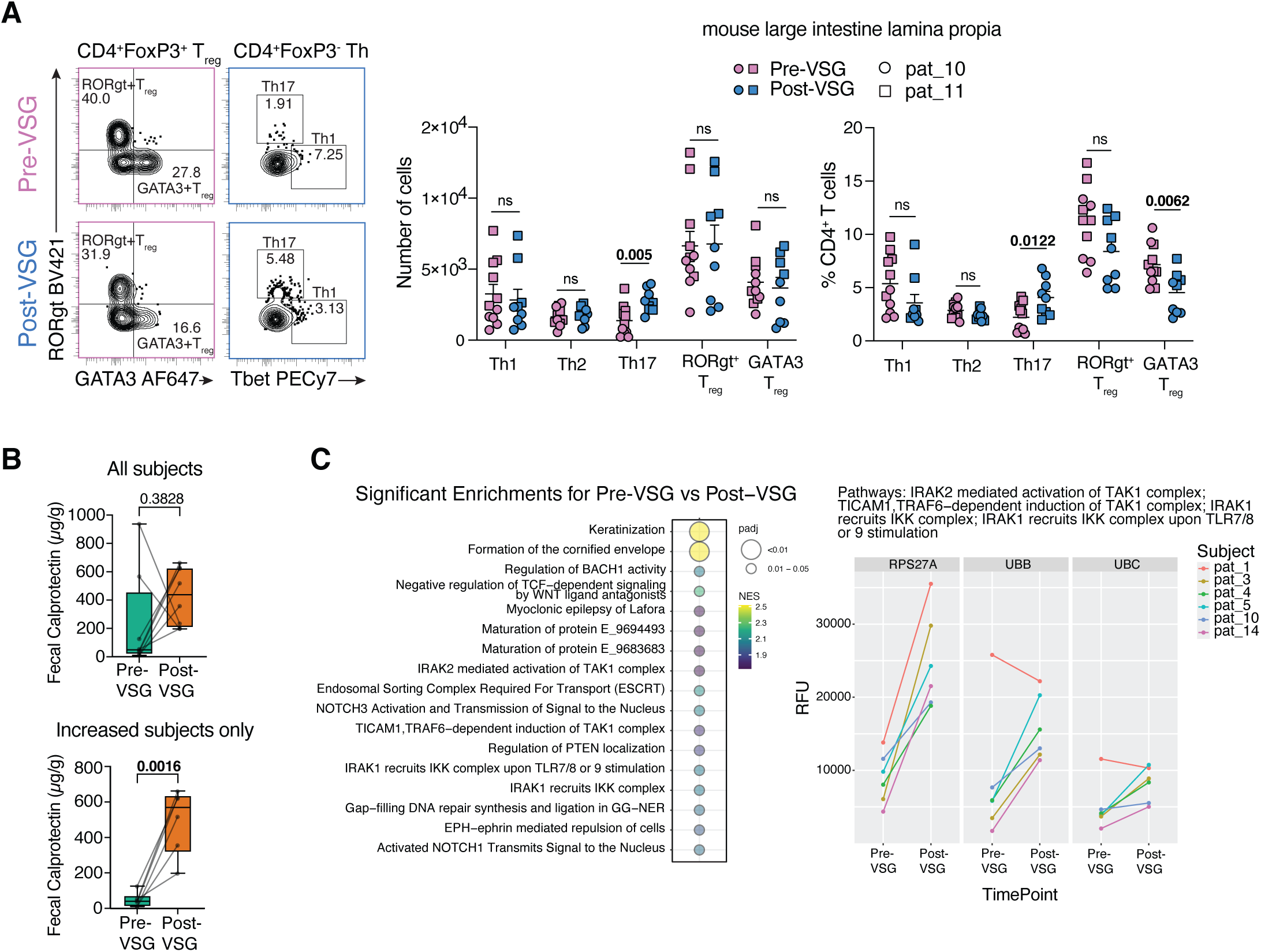
Immune and inflammation assessment in mice and humans. (A) Representative contour plots depicting RORγt+ and GATA3+ Treg populations and Th1 and Th17 populations from the lamina propria cells of the large intestine (left). Gating strategy is shown in SuppFig.6A. The number of live cells for each of the CD4+ T cell subsets and their proportion are plotted (right). Data are based on FMT from 2 participants as shown in the key and are displayed as mean ± standard error of the mean. Statistical significance between the pre-VSG and post-VSG groups was calculated using an unpaired Student’s t-test and non-significance (ns) or p-values are indicated. (B) Fecal calprotectin levels in all human subjects who had paired stool samples available (n=8, top) and all subjects who had an increase in calprotectin with VSG (n=6, bottom). (C) Urine proteomic pathways significantly enriched post-VSG. 1856 proteins quantified in urine were compared between timepoints before and after VSG for 6 subjects and ranked by t-statistic for enrichment analysis using Reactome gene sets. All pathways significantly positively enriched for an increase in change post-VSG are shown (FDR adjusted p < 0.05), with enrichment score and significance indicated by plotted color and size (left). For 4 pathways of interest, expression observed for the 3 leading-edge proteins is shown for all subjects and timepoints measured, where RFU indicates relative fluorescence units (right). Abbreviations: VSG: vertical sleeve gastrectomy.

### FT into germ-free mice

Next, to assess whether the microbiome/metabolome changes seen with VSG had a phenotypic effect, germ-free mice were inoculated with pre-VSG or post-VSG stool from the same participant (Fig.4A). Two human participants were chosen who had both pre-VSG and post-VSG stool stored adequately in glycerol to preserve bacteria viability. Of note, one subject had the second-to-greatest TBWL and the second had prediabetes pre-VSG which resolved post-VSG. Using 16S rRNA gene sequencing, 6 weeks post-FT, mice transplanted with post-VSG stool had a higher alpha diversity (Fig.4B) and different bacterial composition (PERMANOVA Bray Curtis R2 = 0.114, P=0.006, adjusted for human inoculum source, Fig.4C) compared to mice that received pre-VSG stool, which was similar to the human stool microbiota findings. The mouse microbiome samples did also show significant separation based on the human subject used as the inoculum (PERMANOVA Bray Curtis R2=0.495, P=0.001, adjusted for FT timepoint).

### Phenotype changes in germ-free mice

Phenotypic differences resulting from FT with pre-VSG and post-VSG stools were compared. Six weeks post-FT, there was no difference in body weight, food consumption, intraperitoneal glucose tolerance tests, nor tissue weights (epididymal fat, subcutaneous fat and liver weights) between mice that received pre-VSG versus post-VSG stool (Fig.4D-G respectively). Consistently, micro-computed tomography revealed no differences in subcutaneous or intraabdominal adipose tissue volume between the two groups (Fig.4H). There was a significant increase in serum resistin in mice that received post-VSG stool compared to pre-VSG stool (p=0.047), but no differences in other metabolic hormones (GLP-1, insulin, leptin, gastric inhibitory polypeptide, Fig.4I).

### Immune and inflammatory changes in germ-free mice

Immune changes in the murine models were assessed in the large intestine lamina propria and mesenteric lymph nodes using flow cytometry (SuppFig.6A). Of note, mesenteric lymph node data was only available from one set of mice pre- and post-VSG FT. In the large intestine, there was a significant decrease in γδ T cells, a non-significant increase in CD4^+^ T cells and decrease in CD8^+^ T and NK cells in post-VSG mice (Supp.Fig6B). More importantly, there was a significant increase in the number and proportion of Th17 cells, along with a significant decrease in GATA3^+^ regulatory T cell (Treg) proportion in the large intestine and mesenteric lymph nodes of post-VSG mice (Fig.5A, Supp.Fig6C) suggesting an inflammatory phenotype in the individuals studied. Th17 cytokines (IL-17A and IL-22) were also increased in the mesenteric lymph node post-VSG (SuppFig.6D) but did not reach significance in the colon. Sparse Partial Least Squares analysis of the most discriminatory immune and metabolic readouts and microbial taxa showed a distinct separation of pre- and post-VSG parameters indicating a direct or indirect association between the microbiota and observed immune milieu. Notably, the increased Th17 phenotype observed in the post-VSG mice clustered with increases in relative abundances of microbial taxa belonging to Ruminococcaceae, Erysipelatoclostridiceae, and Monoglobus. Conversely, the increased Treg populations in the pre-VSG mice were associated with increased levels of the taxa Parabacteroides (SuppFig.6E).

### Inflammatory changes in humans with VSG

Given the inflammatory phenotype post-VSG observed in the murine model, colonic inflammatory changes were assessed in available human stool samples by fecal calprotectin (FC). 8/12 participants had stool frozen without preservatives to allow for FC assessment. 6/8 individuals had an increase in FC to a clinically elevated level post-VSG (>120ug/g, Fig.5B). Of these 6 participants, the mean FC pre-VSG increased from 47ug/g to 497ug/g post-VSG (p=0.0016, Fig.5B). 2/8 patients with an elevated FC level pre-VSG had a decrease post-VSG. There was no correlation between length of time from surgery and FC levels. Subjects with raised FC post-VSG did not report typical clinical symptoms associated with high levels such as diarrhea. Additionally, the microbiome of subjects with increased FC also exhibited increased post-VSG microbiota similarity compared to pre-VSG where samples were more distinct (p for interaction=0.016, Supplementary Fig.7A-B). No individual taxa correlated with FC increases.

To assess systemic immunity/inflammation changes with VSG, urine samples were utilized due to the unavailability of blood samples. Proteomic analysis was performed using the SomaScan V4.1 Assay, where a subset of 1856 proteins were identified with detectable signals in urine from the overall 7000 analyte panel that has been optimized for peripheral blood (SuppFig.8). When these 1856 proteins were compared between paired pre-VSG and post-VSG urine samples for 6 subjects, no individual proteins differed significantly. However, 17 pathways exhibited significant positive enrichment post-VSG (FDR<0.05, Fig.5C). The top enriched pathways included 4 pathways involved in immune/inflammation regulation: IRAK2 mediated activation of TAK1 complex; TICAM1, TRAF6-dependent induction of TAK1 complex; IRAK1 recruits IKK complex; and IRAK1 recruits IKK complex upon TLR7/8 or 9 stimulation. Enrichment in all 4 pathways was driven by the same set of 3 leading edge proteins related to ubiquitin, and the increased expression observed post-VSG was conserved across participants with one exception (Fig.5C).

## DISCUSSION

We comprehensively examined stool microbiome and metabolome changes pre- and post-VSG in adolescents and assessed if these changes were causal in VSG-associated phenotypes using a murine FT model. Our major findings were 1) increased microbiome diversity post-VSG, with enrichment of several taxa, most notably in those usually associated with the oral cavity; 2) increased ratios of secondary to primary bile acids post-VSG; 3) no differences in metabolic phenotypes in germ-free mice transplanted with pre-VSG and post-VSG stool; 4) an inflammatory phenotype in germ-free mice transplanted with post-VSG stool compared to pre-VSG stool defined by an increase in γδ T cells and Th17 cells and decrease in GATA3^+^ Tregs in the gut and a systemic increase in resistin and 5) a corresponding inflammatory phenotype in a subset of adolescents post-VSG with an increase in FC.

In our cohort, the gut microbiome increased in diversity and changed in composition with VSG. Generally, an increase in alpha diversity is seen with all forms of bariatric surgery in adults, but specific compositional and taxa changes vary by surgery type and study^13^ ^24^. In our study, there was enrichment of taxa post-VSG, most notably in species commonly found in the oral cavity from the *Streptococcus* and *Actinomyces* genera. Some other studies examining microbiome changes post-VSG in adults have also seen an increase in these genera, although species could not be accurately identified due to the use of 16S rRNA sequencing^25^. This increase in oral-associated taxa post-VSG is likely through a variety of mechanisms, including removing some of the physical barrier with VSG that usually prevents the passage of oral taxa and also an increase in stomach pH post-VSG^26^. These changes are very likely due to VSG itself as they are rarely described in weight loss alone^27^. These changes may have potential microbial and immunological consequences. Specifically, post-VSG within the enriched *Streptococcus* species, there was CAZyme enrichment of GH13+CBM 20, a combination shown in fungi to enhance complex starch degradation^28^. Moreover, several ARGs enriched post-VSG were within the enriched *Streptococcus* species. These findings collectively suggest that the *Streptococcus* species that become enriched in the gut microbiome post-VSG are augmented for functions that may enhance their virulence. Overall, the increase in oral-associated taxa may be clinically relevant as there has been increasing recognition that enrichment of oral taxa in the gut maybe associated with several adverse inflammatory patient outcomes, including inflammatory bowel disease (IBD) and subclinical coronary atherosclerosis^29-31^.

There were significant changes in the stool metabolome and microbiome function post-VSG, many of which are considered beneficial. There was a broad increase in the microbiota-driven conversion of host-derived primary bile acids into secondary bile acids post-VSG, suggesting bolstering of the host-to-microbiome communication cycle post-VSG^32^. Further, citrulline, which is understood as both a marker of gut health and to have a role in obesity, was increased post-VSG ^33^ ^34^. This molecule participates in the regulation of numerous pathways relevant to obesity and a therapeutic effect of citrulline supplementation in the regulation of obesity associated metabolic imbalances has also been suggested^35-38^ Moreover, microbially metabolized neurotransmitters including GABA, dopamine, and histamine also trended upward post-VSG in support of a gut-neural homeostasis improvement^39^. Lastly, there was post-VSG enrichment of EC 1.1.1.105, an all-trans retinol dehydrogenase gene, with Vitamin A signaling and homeostasis reported to play a role in mitigating obesity^40^. Overall, it is difficult to assess whether these potentially beneficial trends in fecal metabolites are a result of VSG, post-VSG weight loss or both. The absence of SCFAs changes post-VSG was notable, as changes have previously been reported in an adult study^41^. Large inter-sample variance may have obscured trends in our cohort.

An assessment for a causal role of the microbiome/metabolome changes with VSG performed via FT into germ-free mice did not identify favorable phenotypes such as decreased adiposity or improved glucose tolerance in mice that received post-VSG stool. These findings contrast with similar studies performing human to mouse FTs reporting improvements in body fat, glycemic control and energy expenditure^42-44^ However, those studies involved stool samples from older adults who were prediabetic or diabetic and employed mouse models that differed in age and sex of mice, fecal inoculation, experimental timelines, and diet administered. We may not have seen similar changes as the mice were harvested at 6 weeks following FT, which may not have been long enough to allow phenotypic changes to develop, and only one of the human participants used for FT had prediabetes pre-VSG.

An intriguing finding in our mouse model was the inflammatory phenotype in those receiving post-VSG stools. Given that obesity is considered an inflammatory condition and subjects used for FT experienced weight loss, decreased inflammation in mice receiving post-VSG FT was expected^45^. Indeed, a decrease in different measures of inflammation has been seen following both weight loss of patients with obesity and in one report of decreased gut and systemic inflammation in adults post-VSG^46^ ^47^. Conversely, we observed an increase in the systemic levels of resistin in post-VSG mice, an adipokine increasingly being recognized as playing a role in inflammation^48^ ^49^. Moreover, Th17 cells were increased and GATA3+ Tregs were decreased in the large intestine and mesenteric lymph nodes of post-VSG mice. Numerous studies have described the role of Th17 and imbalance of Th17/Treg ratio in inflammatory bowel disease^50^ ^51^. With relevance to VSG, one study of adults undergoing VSG reported decreases in peripheral Tregs^47^. Further, Erysipelatoclostridiceae that positively correlated with Th17 in our study has been previously associated with inflammation in mouse models^52^ ^53^ In contrast, a recent study in mice demonstrated the beneficial role of microbiota-induced Th17 in protection against diet-induced obesity and metabolic syndrome, a finding not observed in our model^54^. Collectively these data indicate that stool microbiome/metabolome changes from VSG in humans may have gut and systemic inflammatory potential when transplanted to germ-mice, although the underlying mechanisms remain to be elucidated.

When inflammation was assessed in our pediatric participants, FC, a gut specific marker of inflammation, was found to be raised in a subset of adolescents post-VSG, to levels well above the normal reference range. Two studies in adults report persistently raised FC levels after bariatric surgery, with one study showing a persistent increase above baseline 1-year post-VSG.^55^ ^56^. Interestingly in all studies including ours, raised FC levels did not correlate with adverse gastrointestinal symptoms or TBWL. In our study, systemically, 4 pathways involved in immune/inflammation regulation were enriched post-VSG, driven by proteins related to ubiquitin. Ubiquitination of the IKK complex mediated via proteins IRAK and TRAF-6 is an important step in the activation of the NF-kB pathway that plays a critical role in the immune response to microbes including the transcription of pro-inflammatory genes such as IL-2, IL-6, IL-12, TNFα, activation of macrophages and differentiation of CD4^+^ T cells^57^. Increased activation of this pathway has been demonstrated as an important driver of IBD^58^. Furthermore, the gut microbiota has been shown to modulate the expression of this signaling cascade and thus directly influence gut inflammation^59-61^.

Elevated FC levels and enrichment of pathways related to pro-inflammatory responses in humans post-VSG and an increased proportion of intestinal Th17 in comparison to Tregs, with increased resistin levels in mice colonized with post-VSG microbiota, collectively suggest an inflammatory state of subjects post-VSG. This may be clinically relevant given studies, including a large case series and two national database studies, showing an increased incidence of IBD after bariatric surgery^62-64^. One study reported an increased incidence of ulcerative colitis, but not Crohn’s disease, with VSG^63^. It is widely accepted that IBD is at least in part a gut microbiome-mediated disease in susceptible individuals^65^. Therefore, we postulate that the large shifts in the gut microbiome seen post-VSG can have inflammatory potential, and in certain individuals, this may increase their risk of IBD or other inflammatory diseases. It could be argued that increases in FC may be related to the actual VSG rather than gut microbiome changes resulting from VSG. However, against this, is the inflammatory phenotype seen in mice transplanted with post-VSG stool who did not receive surgery themselves and the level of FC post-VSG not correlating with the length of time post-surgery.

Our findings by no means negate the many beneficial effects of VSG in adolescents with obesity. However, they do highlight a new finding that microbiome changes post-VSG may be inflammatory in some adolescents. This paves the way for further research to gain insight into potential inflammatory mechanisms with gut microbiota changes post-VSG. Additionally, larger studies of children and adolescents undergoing VSG followed for a longer period are needed to assess whether microbiome changes and inflammation are persistent, and if those with persistent inflammatory changes are at increased risk for adverse outcomes such as IBD.

Despite this being the first study to our knowledge to comprehensively examine microbiome and metabolome changes post-VSG in adolescents and show that these changes with VSG can be potentially inflammatory, the study has many limitations. The sample size is limited, partly due to the limited number of adolescents undergoing VSG. Additionally, longitudinal samples after 7 months post-VSG were unavailable. While a large increase in oral-associated taxa was seen in the stool post-VSG, no oral samples were available to confirm whether the same taxa from the oral cavity were engrafting in the gut within a specific participant. Additionally, if these oral taxa contribute to the inflammatory phenotype remains to be determined. Finally, a complete assessment of inflammation in adolescents was limited as only stool and urine were available and limited samples were available for the FT murine models to assess whether the findings held across the participant cohort.

In conclusion, large changes in the stool microbiome and metabolome were seen in adolescents post-VSG. There was a notable enrichment of oral-associated taxa in the gut post-VSG. The post-VSG changes in the gut microbiome/metabolome were shown to have inflammatory potential when transferred to a germ-free mouse model. Furthermore, raised FC and inflammatory systemic pathways were seen in a subset of adolescents post-VSG. While VSG is highly effective for weight loss and reduction of comorbidities in adolescents with obesity, we show the novel finding of potential inflammatory microbiome changes post-VSG. This may be of importance given the growing recognition of an increased incidence of IBD after bariatric surgery and warrants further investigation.

## METHODS

### EXPERIMENTAL MODEL AND SUBJECT DETAILS

#### HUMAN METHODS AND ANALYSIS

##### Subjects and samples

Children and adolescents undergoing VSG between January 2021 to February 2023 were enrolled in an Institutional Review Board (IRB) approved longitudinal cohort study “Biorepository of Specimens for Pediatric Obesity Research Use” (Children’s National Hospital IRB protocol #Pro00015976) with signed consent and assent (latter for those >11 years). Inclusion criteria were children and adolescents (≤21 years) undergoing vertical sleeve gastrectomy at the Children’s National Hospital in Washington DC. To be eligible for surgery, children 19 and under required a BMI ≥ 120^th^ percentile of 95th percentile for age and sex with an obesity-related comorbidity and/or BMI ≥ 140^th^ percentile.

Participants were enrolled up to 2 months prior to their planned VSG. After consent was obtained, a stool and urine collection container were mailed to participants. For stool, participants mailed back a sample aliquoted in an OMNIgene-gut tube and OMNImet-gut tube for DNA and metabolite preservation respectively (DNA Genotek Inc)^66^. Wherever possible participants also returned whole stool aliquoted into tubes with glycerol (for bacterial preservation) and whole stool without preservatives. Participants gave a clean catch urine sample mixed with AssayAssure ® Genelock (Sierra Molecular) for protein preservation. All samples were mailed back overnight on icepacks and stored at -80°C until analysis. The same procedure was followed for post-VSG samples. Clinical data was obtained via extraction of medical records from clinical appointments pre-VSG and post-VSG and patient collected data accompanying the samples and was stored in a secure REDCap database. For clinical data analysis, BMI, weight, HbA1c%, LDL, HDL, triglycerides, and ALT passed Shapiro-Wilks normality and were compared pre-VSG and post-VSG via paired t-tests. T2DM, dyslipidemia, and hypertension were compared via McNemar exact tests.

##### Patient involvement

Patients were involved in the design and conduct of the trial. We received input from patients in prior trials for the design of convenient sample collection methods that would minimize inconvenience and were suitable for the planned downstream assays. We intend to disseminate the main results to study participants.

##### Microbiome sequencing

###### DNA Extraction

DNA was extracted from human and murine fecal samples in two stages. First, approximately 50 mg of fecal material and 650 μL MBL lysis buffer from the PowerMicrobiome DNA/RNA EP Kit (Qiagen) were added to Lysis Matrix E (LME) tubes (MP Biomedicals). LME tubes were transferred to a Precelleys 24 Tissue Homogenizer (Bertin Technologies) and fecal samples were homogenized, centrifuged, with the resultant supernatant transferred to a deep-well 96-well plate. The second stage consisted of DNA isolation from the above supernatant using the MagAttract PowerMicrobiome DNA/RNA EP Kit (Qiagen) on an automated liquid handling system as detailed by the manufacturer (Eppendorf).

###### Shotgun Metagenomic Sequencing

Total gene content of the microbiome was assessed through shotgun metagenomic sequencing. Metagenomic libraries were constructed from 100 ng of DNA as starting material using the Illumina DNA Prep kit. Illumina DNA/RNA UD Indexes were used to add sample-specific sequencing indices to both ends of the libraries. An Agilent 4200 TapeStation system with High Sensitivity D5000 ScreenTape (Agilent Technologies, Inc) was used to verify quality and assess final library size. A positive control (MSA-2002 20 Strain Even Mix Whole Cell Material (ATCC)) and a buffer extraction negative control were included. Metagenomic libraries were normalized and pooled at an equimolar concentration. Final pools were diluted to 750 pM and sequenced on a NextSeq2000 sequencer using a paired-end (100x100) NextSeq 1000/2000 P2 (200 cycles) kit (Illumina, Inc).

##### Microbiome analysis

###### Data Processing

The quality of raw paired-end sequence reads was assessed with FastQC^67^ and MutiQC^68^. Adapters revealed by FastQC were removed using bbtools’ bbduk software. Reads then underwent the Whole Genome Sequence Assembly 2 (WGSA2)^69^ protocol using the Nephele platform^70^ (version 2.24.2). In brief, reads were processed with fastp^71^ and minimal trimming and filtering by ensuring an average read quality of 10, a trim of the 3’ end of the read at a quality of 15 and trimming of the 5’ end at a Q score of 20, with additional filtering of reads if they were less than 60 bases after trimming. Decontamination was undertaken using Kraken2^72^ with a database containing the human and mouse genome. After adapter trimming and filtering, samples contained between 15M and 30M paired-end reads, of which between 8.7M and 22M were classified to the bacterial kingdom. Taxonomic identification was performed on the trimmed, error-corrected, and decontaminated reads in Kraken2 with the default RefSeq database.

###### Assembly and Gene Annotation

Within the WGSA2 pipeline, the trimmed, error-corrected and decontaminated reads were assembled into contiguous sequences, or contigs, using metaSPAdes^73^. Reads were recruited back to contigs using bowtie29 and SAMtools^74^ to produce information on scaffold coverage and quality. Protein coding regions (CDS) were predicted from assembled scaffolds using prodigal. Predicted CDS regions were processed by EggNog-mapper2^75^ to identify and annotate genes with KEGG Orthology (KO), Enzyme Commission (EC) and Clusters of Orthologous Genes (COG) identifiers. Abundances were calculated using verse^76^ to obtain Transcripts Per Million (TPM) at the CDS level and summed to obtain TPM by gene.

###### CAZymes

Carbohydrate Active Enzymes (CAZymes) were annotated from assembled metagenomic scaffolds using the dbCAN software^77^ in meta mode with default settings and default databases to obtain eCAMI, HMMER and DIAMOND-based annotations. Annotations were provided at the gene level and merged with predicted gene abundances. Genes were also annotated with taxonomy using Kraken2, creating a table of CAZyme abundances both stratified by taxonomy and unstratified. Gene abundances were normalized to copies per million before analysis. The eCAMI-based CAZyme identification was used in analysis and focused specifically on CAZymes supported by DIAMOND or HMMER when available.

###### Antibiotic Resistance Genes

Antibiotic resistance genes were identified using the Comprehensive Antibiotic Resistance Database with the Resistance Gene Identifier tool v6.0.1, nudging loose hits to strict and including low quality assemblies with prediction of partial genes (ref). Resulting annotations were processed as for CAZymes.

###### Virome

After assembly with WGSA2, assembled scaffolds and binary alignment maps (BAM) were processed for the presence of viral diversity (ssDNA, dsDNA phage, and giant DNA viruses) using Nephele’s DiscoVir pipeline (https://nephele.niaid.nih.gov/pipeline_details/discovir/). Briefly, geNomad predicted viral genomes and fragments using default confidence parameters as defined by the tool, and VERSE calculated read counts of viral genomes based on BAM files^78^ ^79^. Next, CheckV assessed quality and all scaffolds identified as viral were retained for downstream processing and analysis^80^. Viral genomes and fragments greater than 1000 basepairs were clustered with bbtools and MMseqs2 to generated vOTUs which were functionally annotated with DRAM-v with KOfam, Pfam, and Viral Orthologous Group (VOG) databases. Auxillary metabolic genes were identified with VirSorter2.0 and DRAM-v^81-83^. Phage hosts were predicted using iPHoP^84^.

###### Statistical Analysis

All statistical analysis of the microbiome was performed using R 4.3.0. For taxonomic analysis, reads were filtered to those that aligned to the bacterial kingdom and normalized with rarefaction to 8 million reads per sample. Several measures of alpha diversity were calculated, including Chao1 Richness, Observed Taxa, Evenness, Inverse Simpson, and Shannon Diversity using estimate_richness from phyloseq^85^. For all data types, Bray Curtis and Canberra distance matrices were calculated using phyloseq’s distance function. Alpha diversity was compared between pre-VSG and post-VSG samples using linear mixed-effects models within the lmerTest package^86^. The composition of the microbiome was compared against study covariates with Permutational Analysis of Variance (PERMANOVA) using the adonis2 function from the vegan package^87^. Significant relationships were visualized using Principal Coordinates Analysis (PCoA) ordination in phyloseq and ggplot2^88^. All differential abundance analyses were undertaken with Maaslin2^89^, wherein data was analyzed with linear models after log-transformation. Features were filtered if they exhibited a minimum prevalence of less than 10% and a minimum variance of 0.01. When paired samples were included, Subject ID was provided as a random effect. An FDR-corrected p-value less than 0.2 was considered significant.

Co-associated networks of taxa or EC abundances were generated using Weighted Gene Co-Association Network Analysis^90^. In brief, this tool reduces multi-dimensional data to co-associated networks or modules. For taxa-based modules, taxa were included that were found to change nominally after surgery at a false-discovery-corrected p-value of less than 0.5. Blockwise modules were generated using a soft power threshold of 14, an unsigned topology overlap matrix, minimum module size of 20, merge cut height of 0.15, and deepSplit of 3. This process generated eight modules of taxa that were annotated manually by combining the taxonomic information, the average abundance of the taxon across samples, and the module membership, calculated as the correlation of the taxon abundance with the module eigengene.

WGCNA modules were also created for EC abundances using similar settings. ECs were included if they changed significantly due to surgery at an FDR P-value of 0.4. Modules were constructed using the same parameters as above except with a soft power threshold of 7. This resulted in four distinct modules of ECs. These were described using the pathway enrichment tool OmePath^91^, where scores used were based on module membership and calculated as described previously.

Relationships between module eigengenes and metabolites were identified using linear mixed effects models. Results were visualized using the igraph package21 if they were significant at an FDR p-value less than 0.05. Directionality of the results was represented by edge color and data type was indicated through color of nodes.

This work utilized the computational resources of the NIH HPC Biowulf cluster (http://hpc.nih.gov) and the NIAID Locus cluster (http://locus.niaid.nih.gov).

##### Metabolomics

###### Metabolite and Lipid Sample Preparation

For all liquid chromatography mass spectrometry (LCMS) methods, LCMS-grade solvents were used. For bile acid analysis 400 µL of homogenized feces was taken from the fecal collection tubes and added to 500 µL of ice-cold methanol. To each sample 5 µL of the Bile Acid SPLASH® (Avanti Polar Lipids Inc.) and 2 µg of butylated hydroxytoluene was added. Samples were agitated via shaking at 4°C for 20 minutes and then centrifuged at 16k xg for 20 min. An aliquot of the supernatant was taken directly for liquid LCMS analysis.

For short-chain fatty acid (SCFA) and polar metabolomics, a separate 400 µL aliquot of homogenized feces was added to 400 µL of water. Following mixing, 400 µL of chloroform was added. Samples were shaken for 30 minutes at 4°C and subsequently centrifuged at 16k xg for 20 min. 400 µL of the top (aqueous) layer was collected. The aqueous layer was sub-aliquoted for SCFA derivatization or diluted 5x in 50% methanol in water and prepared for LCMS injection.

###### SCFA Derivatization

To preserve SCFAs for analysis an aliquot of the aqueous fraction was derivatized with O-benzylhydroxylamine (O-BHA) as previously described with modifications^92,93^. The reaction buffer contained 1M pyridine and 0.5 M hydrochloric acid in water. To 35 µL of sample, 10 µL of 1M O-BHA in reaction buffer and 10 µL of 1M 1-Ethyl-3-(3-dimethylaminopropyl) carbodiimide in reaction buffer were added. Samples were derivatized for 2 hours at room temperature with constant agitation. Addition of 50 µL of 0.1 % formic acid was used to quench the reaction, which eliminated the potential for formate to be measured in these samples. To extract derivatized molecules 400 µL of ethyl acetate was added. The samples were centrifuged at 16k xg and 4°C for 5 min to induce layering. The upper (organic) layer was collected and dried under vacuum. Samples were resuspended in 300 µL of water for LCMS injection.

###### Liquid Chromatography Mass Spectrometry

Tributylamine and all synthetic molecular references were purchased from Millipore Sigma. LCMS grade water, methanol, isopropanol and acetic acid were purchased through Fisher Scientific.

All samples were separated using a Sciex ExionLC™ AC system and measured using a Sciex 5500 QTRAP® or Sciex 6500+ QTRAP® mass spectrometer.

Polar metabolites were analyzed as previously described^94^. For all metabolomics analysis, quality control samples, consisting of a mixture of the analyzed samples, were injected after every 10 injections to control for signal stability. Samples were analyzed via separate negative ionization and positive ionization methods. For negative mode analysis, a Waters Atlantis T3 column (100 Å, 3 μm, 3 mm × 100 mm) with a gradient from 5 mM tributylamine, 5 mM acetic acid in 2% isopropanol, 5% methanol, 93% water (v/v) to 100% isopropanol over 15 min was used. For positive mode analysis, samples were separated on a Phenomenex Kinetex F5 column (100 Å, 2.6 μm, 2.1 mm × 100 mm) column with a gradient from 100% water with 0.1% formic acid to 95% acetonitrile with 0.1% formic acid over 5 min. Each metabolite was measured using at two distinct multiple reaction monitoring (MRM) signals and a defined retention time.

For SCFA analysis samples were separated on a Waters™ Atlantis dC18 column (100Å, 3 µm, 3 mm X 100 mm) with a 6 min gradient from 5-80 % B with buffer A as 0.1 % formic acid in water and B as 0.1 % formic acid in methanol. All SCFA were measured using positive ionization using MRMs that featured the characteristic 91 daughter ion from O-BHA derivatization. Identity was confirmed via comparison to previous standards.

Bile acid samples were separated on a Phenomenex Kinetex® Polar C18 (100Å, 2.6 µm, 3 mm X 100 mm) using a binary gradient of A: 0.01 % acetic acid in water and B: 0.01 % acetic acid in methanol. A 20 min gradient from 40-100 % B was utilized for separation. Samples were detected in negative MRM mode using previously validated MRMs^95^. Internal bile acid standard signals were used to confirm signal identities and retention times.

###### Metabolomic analysis

All signals were integrated using SciexOS 3.1 (AB Sciex Pte. Ltd.). Signals with greater than 50% missing values were discarded and the remaining missing values were replaced with the lowest registered signal value. All signals with a QC coefficient of variance greater than 30% were discarded. Metabolites with multiple MRMs were quantified with the higher signal-to-noise MRM. Filtered datasets were total sum normalized prior to analysis. The SCFA dataset and the two polar metabolomics datasets were scaled and combined using common signal for serine and succinate for the positive mode metabolite method and the SCFA method respectively. A paired t-test was used for all bile acid and metabolite statistics and a Benjamini-Hochberg method for correction for multiple comparisons was imposed where indicated.

##### Fecal Calprotectin (FC)

FC was assessed using the Buhlmann Fecal Calprotectin ELISA kit (BÜHLMANN fCAL® ELISA, https://buhlmannlabs.com/buhlmann-fcal-elisa/) following the manufacturer’s guidelines. The FCs included all subjects who did not pass Shapiro-Wilks normality, so pre- and post-VSG were compared via a Wilcoxon test. FCs excluding outliers passed normality and were compared pre- and post-VSG via a paired t-test.

##### Urine Proteomics

Urine samples were analyzed using the SomaScan V4.1 Assay, an aptamer-based quantitative proteomic biomarker discovery platform (SomaLogic; Boulder, CO). The assay was run according to manufacturer specifications which includes pH adjustment and buffer exchange by gel filtration prior to normalizing total protein concentration of urine samples to a standard input concentration (https://somalogic.com/wp-content/uploads/2023/09/D0005009_Rev1_2023-09_SomaScan-7K-v4.1-UrinePre-processing-User-Manual.pdf). Data was then subjected to the manufacturer’s standard normalization methods, including adaptive normalization by maximum likelihood by SomaLogic.

Identified enriched gene sets were determined utilizing the pre-ranked gene-set enrichment analysis (GSEA) algorithm, as implemented in the FGSEA R package^96^. Genes were prioritized based on moderated T statistics derived from the limma model’s relevant coefficient, and enrichment analysis was conducted using the Reactome database, with correction of p values applied for multiple sampling. This analysis can be used to identify significant enrichment of a set of foreground genes or proteins, in predefined gene sets, compared against a reference set. Quality control and initial data processing were performed using an R package (https://github.com/foocheung/sqs) and Shiny app^97^.

#### MOUSE METHODS AND ANALYSIS

##### Mice and FT

Germ-free C57BL/6NTac were bred and maintained at the NCI or NIAID Gnotobiotic animal facility. Mice were screened by microbiological culture and 16S rRNA PCR to ensure their germ-free status. Mice were fed a standard irradiated 5KAI diet (LabDiet). Pre-VSG and post-VSG fecal samples from two human subjects stored in 20% glycerol at -80°C were used for fecal microbiota transplant (FMT) into 5-6 week old male mice. Each mouse was orally inoculated with 200 μl of 35-60 mg stool slurry resuspended in sterile pre-reduced PBS in the anaerobic chamber. Stool suspension was stored in a hungate tube. Each mouse was housed in a separate cage in biocontainment racks in the same facility following inoculation. Body weights and feed consumption was measured weekly and stool samples were collected for sequencing. Experiments were performed independently for each human subject and 4-6 mice were inoculated with either pre-VSG or post-VSG stool samples for each subject. All procedures were performed in accordance with approved animal study proposals by NCI or NIAID Animal Care and Use Committees.

##### Glucose tolerance test

Mice were fasted overnight for 12-14 hours. Glucose (2g/kg body weight) was administered intraperitoneally, and glucose measurements were performed via tail bleeds using a glucometer (DSS Precision Xtra) before and at 15, 30, 60, 90, and 120 mins after glucose administration.

##### Micro-computed tomography (Micro-CT)

QuantumGX scanner was used to obtain in-vivo high-resolution micro-CT imaging. Mice were anesthetized using 2.5% isoflurane, eyes protected with Artificial Tears ointment and transferred onto the imaging bed and maintained on 2% isoflurane during the imaging process. Images were obtained in 3 slices at 70 mm magnification and reconstructed. Adipose tissue for each mouse was quantified and analyzed using Analyze14 software. Intra-abdominal and subcutaneous fat were isolated using a threshold range of approximately -300 to -50 Hounsfield units.

##### Tissue harvest and isolation of cells from large intestine and mesenteric lymph nodes

Immediately after micro-CT imaging animals were euthanized with CO_2_ and blood was collected by cardiac exsanguination. Subcutaneous fat from the abdominal region, epididymal fat pads, and liver were harvested and weighed. Mesenteric lymph nodes were harvested and cells were isolated by passing through a 70-μm cell strainer, centrifuged at 1500 rpm for 5 mins for flow cytometry analysis. Large intestines (LI, cecum and colon) were collected and placed on ice-cold complete media (RPMI 1640 supplemented with 20mM HEPES, 2mM L-glutamine, 1mM sodium pyruvate, 1mM nonessential amino acids, 50 mM β-mercaptoethanol, 100U/ml penicillin and 100 mg/ml streptomycin) + 3% fetal bovine serum (FBS). The tissue was then opened longitudinally, fecal contents removed and cut into 1-2 cm pieces, and incubated in 20 ml of complete media + 3% FBS + 5mM EDTA + 0.145 mg/ml DL-dithiothreitol for 20 mins at 37°C and 5% CO^2^ with shaking. To remove epithelial cells, LI were strained and vigorously shaken in a 50 ml tube containing 10 ml complete media + 2mM EDTA, thrice. LI were then finely chopped and digested with 10 ml of digestion media (complete media + 0.1 mg/ml Liberase TL, Roche + 0.05% DNase I, Sigma) for 25 mins at 37°C and 5% CO^2^ with shaking. The digestion was stopped by adding 10 ml complete media + 3% FBS. Digested tissues were then passed through 100-μm cell strainers, centrifuged at 1500 rpm for 5 mins, followed by straining through 40-μm cell strainers and centrifugation at 1500 rpm for 5 mins. Isolated lamina propria cells were then resuspended in complete media + 10% FBS for flow cytometry analysis.

##### Spectral flow cytometry

Isolated single-cell suspensions were assessed for cytokine production potential by stimulation with Cell Stimulation cocktail 500X (Thermo Fisher Scientific) prepared in complete media + 10% FBS for 2.5 hours at 37°C. To assess lymphoid cells, cells were incubated with Zombie NIR Fixable Viability Dye for 15 mins at room temperature. Cells were then washed and incubated with a cocktail of fluorescently labeled antibodies prepared in PBS + 1% FBS +10% Brilliant Stain Buffer + 10% TruStain FcX for 25 mins in the dark at 4°C: anti-CD45, anti-TCRβ, anti-CD44, anti-CD90.2, anti-CD8β, anti-CD4, anti-TCRγδ and anti-NK1.1. Cells were then fixed/permeabilized in eBioscience FoxP3 Fixation/Permeabilization Solution kit overnight at 4°C and stained with a cocktail of fluorescently labelled antibodies against intracellular antigens prepared in eBioscience intracellular staining buffer: anti-FoxP3, anti-GATA3, anti-RORγt, anti-Tbet, anti-IFNγ, anti-IL17A and anti-IL22.

All samples were collected on an Aurora spectral cytometer (Cytek). Spectral unmixing was performed using single-control controls using cells from corresponding tissues or UltraComp eBeads (Invitrogen) and data were analyzed using FlowJo version 10.

##### Serum metabolic hormones measurement

Mouse serum concentrations of metabolic hormones were assessed using the Hormone Exp Panel kit (Millipore, MMHE-44K-06) according to the manufacturer’s protocol and measured using Luminex MAGPIX Instrument (Bio-Rad). Data were analyzed using GraphPad Prism9.0 (Graph Pad software, La Jolla, CA, USA) with unpaired t-tests. Differences were considered to be statistically significant when p<0.05.

##### Microbiome sequencing:16S ribosomal RNA Gene Sequencing

DNA extraction was as per human samples. Microbiome composition was assessed via dual-index amplification of the V4 region of the 16S ribosomal RNA gene (16S rRNA). This method used the V4 16S rRNA 515F and 806R primers with individual sample-specific indices and Illumina sequencing adapters appended as previously described^98^. The V4 region was amplified using: 5 μM of F/R primers, 1X Phusion High-Fidelity DNA Polymerase (New England Biolabs) and 100 ng of DNA as starting material. PCR conditions for amplification were as follows: initial template denaturation at 98°C for 60s; 25 cycles of denaturation at 98°C for 10s, primer annealing 55°C for 30s, and template extension at 72°C 60s; with a final template extension at 72°C for 5 min. AMPure XP beads (Beckman-Coulter) at a 1:1 ratio with the above PCR reaction were used to isolate final PCR products.

Final 16S rRNA V4 libraries were quantified using the KAPA qPCR Library Quantification Kit (Kapa Biosystems) and pooled at an equimolar concentration. Pools were normalized to 8 pM, spiked-in with 15% phiX control library (Illumina) to add sequence diversity, and sequenced on the Illumina MiSeq instrument utilizing the 600 cycle Paired-End (250x250) Reagent Plate with the addition of 16S V4 rRNA specific sequencing primers^98^.

##### Microbiome analysis

Samples collected from the murine study at six weeks as well as the human inoculum (pre-VSG and post-VSG samples from two subjects) underwent 16S rRNA sequencing on the Illumina MiSeq. The resulting sequences were reviewed for quality using FastQC and multiQC through Nephele’s microbiome analysis platform. Sequences were trimmed to 210 bases on the forward reads and 180 bases on the reverse read, and those with a maximum expected error greater than two were filtered out, reads then underwent Divisive Amplicon Denoising Algorithm 2 (DADA2) utilizing Nephele’s microbiome analysis platform^99^. After the identification of sequence variants through denoising, they were checked for chimeras and assigned taxonomy up to the species level using the SILVA database. If a sequence variant aligned to multiple species with 100% identity, all species were listed. Once reads were agglomerated into a table of counts, reads from negative controls were subtracted from samples to be conservative about potential sources of contamination.

The sequence variant table was rarefied to 70,000 reads per sample after reviewing rarefaction curves and sequences available per sample. Alpha diversity metrics, including Chao1 richness, Evenness, Inverse Simpson and Shannon diversity values, as well as a Bray Curtis distance matrix, were calculated using phyloseq. Linear models determined differences in alpha diversity, and vegan’s PERMANOVA utilizing marginal adjustment for the subject that served as the inoculum source identified differences in community composition between the gut microbiome of pre-VSG and post-VSG mice.

##### Multi-Omic analysis

DIABLO from the mixOmics package implemented sparse Partial Least Squares (sPLS) to perform discriminant analysis of multi-omic data^100^. Extensive k-fold cross validation and leave-one-out analysis was performed but did not provide stable estimates to obtain an optimal number of features. Therefore, the top 10 most discriminant features from each data type and human inoculum sources that differentiated pre-VSG mice from post-VSG mice were selected for inclusion in the combined visualization of microbiota and flow data sources.

## Data Availability

All data generated or analyzed during this study are included in this published article and its online supplemental information files.
Sequencing reads for human and murine analyses are deposited in the Sequence Read Archive (SRA) under accession number PRJNA1093424 at the following reviewer link: https://dataview.ncbi.nlm.nih.gov/object/PRJNA1093424?reviewer=gbe14pmjol31cvc64bn6vh4f4r.

## FIGURE LEGENDS

**SuppFig.1. Pre-VSG, microbiome diversity and composition related to Type 2 Diabetes status differed, but resolved post-VSG**. (A) Subjects with established diabetes exhibited significantly distinct bacterial microbiota diversity (P < 0.05). (B) Microbiota composition was also highly distinct between subjects with established T2DM and those without (Bray Curtis PERMANOVA R^2^=0.44, P = 0.001). Abbreviations: VSG: vertical sleeve gastrectomy, T2DM: type 2 diabetes mellitus, PERMANOVA: Permutational Analysis of Variance.

**SuppFig.2. Enzyme Commission genes found to be differentially abundant post-VSG formed three distinct modules of co-associated genes.** (A) PCoA Plot of Enzyme Commission composition using Canberra distance. R^2^ and p-values are derived from PERMANOVA. (B) Differentially abundant enzyme class post-VSG. Enzyme class to the left decrease post-VSG and enzymes to the right increase post-VSG. (C) Hierarchical clustering representation of WGCNA modules based on moderately differential (FDR P < 0.4) pre- and post-VSG. Identified modules are represented by their color underneath the dendrogram. (D) All modules exhibited increases in abundance post-VSG (LME P <0.01). (E) Genera in the microbiome that contribute to each of the three differentially abundant networks described in panels (C) and (D). Abbreviations: VSG: vertical sleeve gastrectomy, FDR: False Discovery Rate, LME: Linear Mixed Effects, PCoA: Principal Coordinates Analysis, PERMANOVA: Permutational Analysis of Variance, WGCNA: Weighted Gene Co-Association Network Analysis.

**SuppFig.3. Metagenomic Viral Diversity and composition is not impacted by vertical sleeve gastrectomy.** (A) Identified viruses were primarily comprised of Caudoviricetes. Donut plot represents abundances across both pre-VSG (left) and post-VSG (right) samples for each taxonomic category. (B) Host taxonomy of identified viruses including *Bacteroides*, *Bacteroidaceae*, and *Prevotella,* as well as several with no known hosts. As in (A), plot represents summarized abundances across both pre-VSG (left) and post-VSG (right) samples for each viral host. (C) Viral taxonomic diversity does not change due to VSG with respect to either Chao1 richness or evenness. (D) Viral taxonomic composition does not change signficantly due to VSG. Exploration of either (E) viral Protein Family (PFAM) diversity or (F) composition did not reveal significant differences due to VSG. Points indicate individual samples and lines connect paired samples. P-values for diversity were generated from a linear mixed-effects model with the subject as the random effect. Ellipses in composition plots represent 20% confidence intervals for each group. PERMANOVA calculated the R^2^ and p-values with the subject as the strata variable. Abbreviations: VSG: vertical sleeve gastrectomy, PERMANOVA (Permutational Analysis of Variance), PFAM (Protein Family).

**SuppFig.4.** Metabolites (circles) correlate with clinical variables (squares) (A) prior to and (B) after VSG (p < 0.05). Pink lines indicate positive associations between metabolites and clinical variables while blue lines indicate negative associations. Relationships were identified with non-parametric statistics (Wilcox Rank-sum for binary variables and Spearman correlation for continuous variables). Abbreviations: VSG: vertical sleeve gastrectomy, AA: Amino Acid, FA-Car: Fatty Acid Carnitines, TCA: Tricarboxylic Acid, NT: Neurotransmitter, SCFA: Short Chain Fatty Acid.

**SuppFig.5. Metabolites change in concert with networks of taxa.** (A) Taxa exhibiting at least moderate change after VSG (P FDR < 0.5) formed several co-associated networks using Weighted Gene Co-Association Network Analysis (WGCNA). (B) These networks were characterized by specific taxa that collectively changed in abundance. P-values were generated from linear mixed effects models. (C) These differential networks changed in concert with several changing metabolites. Each line indicates a significant Spearman correlation between the change of metabolites (difference in value before and after) and the change in microbial taxa (difference in mean abundance before and after). Positive correlations are indicated in pink and negative correlations are indicated in blue. The weight of the line indicates the strength of the correlation. Modules identified in panels (A) and (B) are noted by their characteristic taxa. Abbreviations: VSG: vertical sleeve gastrectomy, AA: Amino Acid, FA-Car: Fatty Acid Carnitine, TCA: Tricarboxylic Acid, NT: Neurotransmitter, SCFA: Short Chain Fatty Acid.

**SuppFig.6. Additional flow cytometry data of mice receiving FT with pre-VSG and post-VSG stool**. (A) Gating strategy for lymphoid cells is shown. (B) Number of live lymphoid cells in the large intestine lamina propria and mesenteric lymph node are plotted. (C-D). Number of live cells for each of the CD4^+^ T cell subsets, their proportion in the mesenteric lymph node (C) and proportion of cytokine producing CD4^+^ T cells (D) are plotted. Groups and participants are identified as shown in the key in (B). Data are displayed as mean ± standard error of the mean. Statistical significance between pre-VSG and post-VSG groups was calculated using unpaired Student’s t-test and non-significance (ns) or p-values are indicated. (E) Top 10 Amplicon sequence variants and flow cytometry features that discriminate pre-VSG vs post-VSG samples from each subject used for FT (n=36 features) identified using mixOmics; rows are z-score scaled. Abbreviations: FT: fecal transplant, VSG: vertical sleeve gastrectomy.

**SuppFig.7. (**A) PCoA plot representing the 6 subjects with increasing calprotectin values post-VSG, in which post-VSG samples become more related to each other. PERMANOVA calculated the interaction between before-vs-after VSG samples (time) and calprotectin. (B) Paired Distance within Pre-VSG samples, within Post-VSG samples, and between Pre-VSG and Post-VSG samples, showing that distance within Pre-VSG samples exhibit higher distance than distances within Post-VSG samples. P-values calculated with Wilcoxon Rank-Sum Test.

**SuppFig.8.** Validation of urine proteomics against serum proteomics in a separate pediatric cohort. The SomaScan Assay utilizes 7000 SOMAmer Reagents optimized for detection of proteins in peripheral blood serum or plasma. To determine the subset that could be measured in urine, SOMAmer Reagents were classified by comparing their average signal in a cohort of 53 urine samples to background and saturation thresholds. SOMAmer Reagents with a signal-to-noise ratio <1.7 (n=5705) or a saturating signal of RFU >80000 (n=35) were excluded. 1856 SOMAmer Reagents remained within the detectable range of the assay when using urine in this cohort of samples.

**Supp Table 1**. Excel file containing taxa enriched post-VSG, related to Figure 1.

**Supp Table 2**. Excel file containing CAZymes enriched post-VSG, post-VSG CAZymes related to Total Body Weight Loss, Antibiotic Resistance Genes enriched post-VSG, related to Figure 2.

**Supp Table 3**. Excel file containing Enzyme Commission genes enriched post-VSG and pathway enrichments of EC networks, related to Supplemental Figure 2.

**Supp Table 4**. Excel file containing pre-VSG and post-VSG correlations between metabolites and clinical metadata, related to Supplemental Figure 4.

**Supp Table 5**. Excel file containing module assignments of bacterial taxa and correlations between bacterial taxa module changes and metabolite changes, related to Supplemental Figure 5.

## DATA AVAILIBILITY STATEMENT

All data generated or analyzed during this study are included in this published article and its online supplemental information files.

Sequencing reads for human and murine analyses are deposited in the Sequence Read Archive (SRA) under accession number PRJNA1093424 at the following reviewer link: https://dataview.ncbi.nlm.nih.gov/object/PRJNA1093424?reviewer=gbe14pmjol31cvc64bn6vh4f4r.

Metabolite data is deposited at https://figshare.com/articles/dataset/Supporting_data_for_bariatric_surgery_patient_fecal_metabolomics_and_bile_acid_measurements/25864327.

This paper does not report original code.

## FUNDING

This work was supported by the National Institute of Allergy and Infectious Diseases (NIAID) of the National Institutes of Health under the Division of Intramural Research, NIAID, NIH (Hourigan), BCBB Support Services Contract HHSN316201300006W/75N93022F00001 to Medical Science & Computing (McCauley, Subramanian, McCauley), the Thrasher Research Fund Early Career Award #01484 (Akagbosu), and the American Pediatric Association Research in Academic Pediatrics Initiative on Diversity (RAPID) grant with funding from the National Institute of Diabetes and Digestive and Kidney Diseases (NIDDK) Grant R25DK096944 (Akagbosu). The funders had no role in the design and conduct of the study; collection, management, analysis, and interpretation of the data; preparation, review, or approval of the manuscript; and decision to submit the manuscript for publication. The content is solely the responsibility of the authors and does not necessarily represent the official views of the National Institutes of Health.

The authors acknowledge the NCI Gnotobiotic Animal Facility Staff for their assistance in performing the germ-free mice experiments. This work used the Office of Cyber Infrastructure and Computational Biology High Performance Computing cluster at NIAID, Bethesda, MD and the Center for Genetic Medicine Research in the Children’s National Research Institute under Dr. Robert Freishtat.

## AUTHOR CONTRIBUTIONS

Cynthia O Akagbosu, Kathryn McCauley and Sivaranjani Namasivayam contributed equally to this paper (first authors).

Evan P Nadler and Suchitra K Hourigan contributed equally to this paper (senior authors).

C.O.A., S.N., E.P.N. and S.K.H. conceived and designed the project. All authors acquired, analyzed, or interpreted data from the study. K.E.M., S.N., H.N.R-S., M.B., B.S., Q.C. and F.C. performed statistical analysis. C.O.A and S.K.H. obtained funding for the project. E.P.N and S.K.H supervised the project. C.O.A., K.M., S.N., B.S. and S.K.H. wrote the original draft. All authors reviewed and edited the manuscript, approved the final submitted manuscript and have agreed both to be personally accountable for the author’s own contributions and to ensure that questions related to the accuracy or integrity of any part of the work, even ones in which the author was not personally involved, are appropriately investigated, resolved, and the resolution documented in the literature.

## ETHICS DECLARATIONS

### Ethics approval and consent to participate

The study was approved by an Institutional Review Board (Children’s National Hospital IRB protocol #Pro00015976) and all participants provided written informed consent ± assent to participate.

### Declaration of interests

B.A.S. is a former SomaLogic, Inc. (Boulder, CO, USA) employee and a company shareholder.

All other authors declare that they have no competing interests.

## ABBREVIATIONS

VSG: Vertical Sleeve Gastrectomy
T2DM: Type 2 Diabetes Mellitus
MASLD: metabolic dysfunction-associated steatotic liver disease
FT: Fecal transplant
GLP-1: Glucagon-like peptide-1
BMI: Body mass index
TBWL: Total body weight loss
ALT: Alanine aminotransferase
LDL: Low-density lipoprotein
HDL: High-density lipoprotein
CAZymes: Carbohydrate-Active Enzymes
ARG: Antibiotic resistance genes
EC: Enzyme Commission
SCFA: Short-chain fatty acids
HbA1c: Hemoglobin A1C
FC: Fecal Calprotectin
IBD: Inflammatory Bowel Disease

## Notes

### Author Declarations

Children's National Hospital IRB have ethical approval for this work.

## REFERENCES

1. National Institute of Diabetes and Digestive and Kidney Diseases Overweight & Obesity Statistics|NIDDK 2021 [Available from: https://www.niddk.nih.gov/health-information/health-statistics/overweight-obesity accessed May 24 2024.

2. Andes LJ, Cheng YJ, Rolka DB, et al. Prevalence of Prediabetes Among Adolescents and Young Adults in the United States, 2005-2016. JAMA Pediatr 2020;174(2):e194498. doi: 10.1001/jamapediatrics.2019.4498 [published Online First: 20200203]

3. Simmonds M, Llewellyn A, Owen CG, et al. Predicting adult obesity from childhood obesity: a systematic review and meta-analysis. Obes Rev 2016;17(2):95–107. doi: 10.1111/obr.12334 [published Online First: 20151223]

4. Juonala M, Magnussen CG, Berenson GS, et al. Childhood adiposity, adult adiposity, and cardiovascular risk factors. N Engl J Med 2011;365(20):1876–85. doi: 10.1056/NEJMoa1010112

5. Inge TH, Courcoulas AP, Jenkins TM, et al. Weight Loss and Health Status 3 Years after Bariatric Surgery in Adolescents. N Engl J Med 2016;374(2):113–23. doi: 10.1056/NEJMoa1506699 [published Online First: 20151106]

6. Pratt JSA, Browne A, Browne NT, et al. ASMBS pediatric metabolic and bariatric surgery guidelines, 2018. Surg Obes Relat Dis 2018;14(7):882-901. doi: 10.1016/j.soard.2018.03.019 [published Online First: 20180323]

7. Hampl SE, Hassink SG, Skinner AC, et al. Clinical Practice Guideline for the Evaluation and Treatment of Children and Adolescents With Obesity. Pediatrics 2023;151(2) doi: 10.1542/peds.2022-060640

8. Steinberger AE, Nickel KB, Keller M, et al. National Trends in Pediatric Metabolic and Bariatric Surgery: 2010-2017. Pediatrics 2022;150(6) doi: 10.1542/peds.2022-057316

9. McCarty TR, Jirapinyo P, Thompson CC. Effect of Sleeve Gastrectomy on Ghrelin, GLP-1, PYY, and GIP Gut Hormones: A Systematic Review and Meta-analysis. Ann Surg 2020;272(1):72-80. doi: 10.1097/sla.0000000000003614

10. Crovesy L, Masterson D, Rosado EL. Profile of the gut microbiota of adults with obesity: a systematic review. Eur J Clin Nutr 2020;74(9):1251–62. doi: 10.1038/s41430-020-0607-6 [published Online First: 20200330]

11. Ridaura VK, Faith JJ, Rey FE, et al. Gut microbiota from twins discordant for obesity modulate metabolism in mice. Science 2013;341(6150):1241214. doi: 10.1126/science.1241214

12. Hamamah S, Hajnal A, Covasa M. Influence of Bariatric Surgery on Gut Microbiota Composition and Its Implication on Brain and Peripheral Targets. Nutrients 2024;16(7) doi: 10.3390/nu16071071 [published Online First: 20240405]

13. Davies NK, O’Sullivan JM, Plank LD, et al. Altered gut microbiome after bariatric surgery and its association with metabolic benefits: A systematic review. Surg Obes Relat Dis 2019;15(4):656–65. doi: 10.1016/j.soard.2019.01.033 [published Online First: 20190207]

14. Akagbosu CO, Nadler EP, Levy S, et al. The Role of the Gut Microbiome in Pediatric Obesity and Bariatric Surgery. Int J Mol Sci 2022;23(23) doi: 10.3390/ijms232315421 [published Online First: 20221206]

15. Yatsunenko T, Rey FE, Manary MJ, et al. Human gut microbiome viewed across age and geography. Nature 2012;486(7402):222-7. doi: 10.1038/nature11053 [published Online First: 2012/06/16]

16. Hollister EB, Riehle K, Luna RA, et al. Structure and function of the healthy pre-adolescent pediatric gut microbiome. Microbiome 2015;3:36. doi: 10.1186/s40168-015-0101-x [published Online First: 20150826]

17. Agans R, Rigsbee L, Kenche H, et al. Distal gut microbiota of adolescent children is different from that of adults. FEMS Microbiol Ecol 2011;77(2):404–12. doi: 10.1111/j.1574-6941.2011.01120.x [published Online First: 20110601]

18. Shelton CD, Sing E, Mo J, et al. An early-life microbiota metabolite protects against obesity by regulating intestinal lipid metabolism. Cell Host Microbe 2023;31(10):1604–19.e10. doi: 10.1016/j.chom.2023.09.002 [published Online First: 20231003]

19. Vu K, Lou W, Tun HM, et al. From Birth to Overweight and Atopic Disease: Multiple and Common Pathways of the Infant Gut Microbiome. Gastroenterology 2021;160(1):128–44.e10. doi: 10.1053/j.gastro.2020.08.053 [published Online First: 2020/09/19]

20. Szczudlik E, Stępniewska A, Bik-Multanowski M, et al. The age of the obesity onset is a very important factor for the development of metabolic complications and cardiovascular risk in children and adolescents with severe obesity. Eur J Pediatr 2024 doi: 10.1007/s00431-024-05636-x [published Online First: 20240615]

21. Draijer L, Voorhoeve M, Troelstra M, et al. A natural history study of paediatric non-alcoholic fatty liver disease over 10 years. JHEP Rep 2023;5(5):100685. doi: 10.1016/j.jhepr.2023.100685 [published Online First: 20230125]

22. Tommerdahl KL, Kendrick J, Nelson RG, et al. Youth versus adult-onset type 2 diabetic kidney disease: Insights into currently known structural differences and the potential underlying mechanisms. Clin Sci (Lond) 2022;136(21):1471–83. doi: 10.1042/cs20210627

23. Wardman JF, Bains RK, Rahfeld P, et al. Carbohydrate-active enzymes (CAZymes) in the gut microbiome. Nat Rev Microbiol 2022;20(9):542–56. doi: 10.1038/s41579-022-00712-1 [published Online First: 20220328]

24. Farin W, Oñate FP, Plassais J, et al. Impact of laparoscopic Roux-en-Y gastric bypass and sleeve gastrectomy on gut microbiota: a metagenomic comparative analysis. Surg Obes Relat Dis 2020;16(7):852–62. doi: 10.1016/j.soard.2020.03.014 [published Online First: 20200320]

25. Paganelli FL, Luyer M, Hazelbag CM, et al. Roux-Y Gastric Bypass and Sleeve Gastrectomy directly change gut microbiota composition independent of surgery type. Sci Rep 2019;9(1):10979. doi: 10.1038/s41598-019-47332-z [published Online First: 20190729]

26. Porat D, Vaynshtein J, Gibori R, et al. Stomach pH before vs. after different bariatric surgery procedures: Clinical implications for drug delivery. Eur J Pharm Biopharm 2021;160:152–57. doi: 10.1016/j.ejpb.2021.01.016 [published Online First: 20210130]

27. Koutoukidis DA, Jebb SA, Zimmerman M, et al. The association of weight loss with changes in the gut microbiota diversity, composition, and intestinal permeability: a systematic review and meta-analysis. Gut Microbes 2022;14(1):2020068. doi: 10.1080/19490976.2021.2020068

28. Sidar A, Voshol GP, Vijgenboom E, et al. Novel Design of an α-Amylase with an N-Terminal CBM20 in Aspergillus niger Improves Binding and Processing of a Broad Range of Starches. Molecules 2023;28(13) doi: 10.3390/molecules28135033 [published Online First: 20230627]

29. Liao C, Rolling T, Djukovic A, et al. Oral bacteria relative abundance in faeces increases due to gut microbiota depletion and is linked with patient outcomes. Nat Microbiol 2024 doi: 10.1038/s41564-024-01680-3 [published Online First: 20240502]

30. Read E, Curtis MA, Neves JF. The role of oral bacteria in inflammatory bowel disease. Nat Rev Gastroenterol Hepatol 2021;18(10):731–42. doi: 10.1038/s41575-021-00488-4 [published Online First: 20210816]

31. Sayols-Baixeras S, Dekkers KF, Baldanzi G, et al. Streptococcus Species Abundance in the Gut Is Linked to Subclinical Coronary Atherosclerosis in 8973 Participants From the SCAPIS Cohort. Circulation 2023;148(6):459–72. doi: 10.1161/circulationaha.123.063914 [published Online First: 20230712]

32. Wahlström A, Sayin SI, Marschall HU, et al. Intestinal Crosstalk between Bile Acids and Microbiota and Its Impact on Host Metabolism. Cell Metab 2016;24(1):41–50. doi: 10.1016/j.cmet.2016.05.005 [published Online First: 20160616]

33. Fragkos KC, Forbes A. Citrulline as a marker of intestinal function and absorption in clinical settings: A systematic review and meta-analysis. United European Gastroenterol J 2018;6(2):181–91. doi: 10.1177/2050640617737632 [published Online First: 20171012]

34. Crenn P, Messing B, Cynober L. Citrulline as a biomarker of intestinal failure due to enterocyte mass reduction. Clin Nutr 2008;27(3):328–39. doi: 10.1016/j.clnu.2008.02.005 [published Online First: 20080428]

35. Azizi S, Mahdavi R, Mobasseri M, et al. The impact of L-citrulline supplementation on glucose homeostasis, lipid profile, and some inflammatory factors in overweight and obese patients with type 2 diabetes: A double-blind randomized placebo-controlled trial. Phytother Res 2021;35(6):3157–66. doi: 10.1002/ptr.6997 [published Online First: 20210420]

36. Eshreif A, Al Batran R, Jamieson KL, et al. l-Citrulline supplementation improves glucose and exercise tolerance in obese male mice. Exp Physiol 2020;105(2):270–81. doi: 10.1113/ep088109 [published Online First: 20200115]

37. Holguin F, Grasemann H, Sharma S, et al. L-Citrulline increases nitric oxide and improves control in obese asthmatics. JCI Insight 2019;4(24) doi: 10.1172/jci.insight.131733 [published Online First: 20191219]

38. Joffin N, Jaubert AM, Durant S, et al. Citrulline induces fatty acid release selectively in visceral adipose tissue from old rats. Mol Nutr Food Res 2014;58(9):1765–75. doi: 10.1002/mnfr.201400053 [published Online First: 20140610]

39. Strandwitz P. Neurotransmitter modulation by the gut microbiota. Brain Res 2018;1693(Pt B):128–33. doi: 10.1016/j.brainres.2018.03.015

40. Blaner WS. Vitamin A signaling and homeostasis in obesity, diabetes, and metabolic disorders. Pharmacol Ther 2019;197:153–78. doi: 10.1016/j.pharmthera.2019.01.006 [published Online First: 20190129]

41. Juárez-Fernández M, Román-Sagüillo S, Porras D, et al. Long-Term Effects of Bariatric Surgery on Gut Microbiota Composition and Faecal Metabolome Related to Obesity Remission. Nutrients 2021;13(8) doi: 10.3390/nu13082519 [published Online First: 20210723]

42. Anhê FF, Zlitni S, Zhang SY, et al. Human gut microbiota after bariatric surgery alters intestinal morphology and glucose absorption in mice independently of obesity. Gut 2023;72(3):460–71. doi: 10.1136/gutjnl-2022-328185 [published Online First: 20220825]

43. Yadav J, Liang T, Qin T, et al. Gut microbiome modified by bariatric surgery improves insulin sensitivity and correlates with increased brown fat activity and energy expenditure. Cell Rep Med 2023;4(5):101051. doi: 10.1016/j.xcrm.2023.101051

44. Tremaroli V, Karlsson F, Werling M, et al. Roux-en-Y Gastric Bypass and Vertical Banded Gastroplasty Induce Long-Term Changes on the Human Gut Microbiome Contributing to Fat Mass Regulation. Cell Metab 2015;22(2):228–38. doi: 10.1016/j.cmet.2015.07.009

45. Rohm TV, Meier DT, Olefsky JM, et al. Inflammation in obesity, diabetes, and related disorders. Immunity 2022;55(1):31–55. doi: 10.1016/j.immuni.2021.12.013

46. Bianchi VE. Weight loss is a critical factor to reduce inflammation. Clin Nutr ESPEN 2018;28:21–35. doi: 10.1016/j.clnesp.2018.08.007 [published Online First: 20180903]

47. Fukuda N, Ojima T, Hayata K, et al. Laparoscopic sleeve gastrectomy for morbid obesity improves gut microbiota balance, increases colonic mucosal-associated invariant T cells and decreases circulating regulatory T cells. Surg Endosc 2022;36(10):7312–24. doi: 10.1007/s00464-022-09122-z [published Online First: 20220219]

48. Pang SS, Le YY. Role of resistin in inflammation and inflammation-related diseases. Cell Mol Immunol 2006;3(1):29–34.

49. Taouis M, Benomar Y. Is resistin the master link between inflammation and inflammation-related chronic diseases? Mol Cell Endocrinol 2021;533:111341. doi: 10.1016/j.mce.2021.111341 [published Online First: 20210531]

50. Schnell A, Littman DR, Kuchroo VK. T(H)17 cell heterogeneity and its role in tissue inflammation. Nat Immunol 2023;24(1):19–29. doi: 10.1038/s41590-022-01387-9 [published Online First: 20230103]

51. Chen L, Ruan G, Cheng Y, et al. The role of Th17 cells in inflammatory bowel disease and the research progress. Front Immunol 2022;13:1055914. doi: 10.3389/fimmu.2022.1055914 [published Online First: 20230109]

52. Kaakoush NO. Insights into the Role of Erysipelotrichaceae in the Human Host. Front Cell Infect Microbiol 2015;5:84. doi: 10.3389/fcimb.2015.00084 [published Online First: 20151120]

53. Miyauchi E, Kim SW, Suda W, et al. Gut microorganisms act together to exacerbate inflammation in spinal cords. Nature 2020;585(7823):102-06. doi: 10.1038/s41586-020-2634-9 [published Online First: 20200826]

54. Kawano Y, Edwards M, Huang Y, et al. Microbiota imbalance induced by dietary sugar disrupts immune-mediated protection from metabolic syndrome. Cell 2022;185(19):3501–19.e20. doi: 10.1016/j.cell.2022.08.005 [published Online First: 20220829]

55. Westerink F, Huibregtse I, De Hoog M, et al. Faecal Inflammatory Biomarkers and Gastrointestinal Symptoms after Bariatric Surgery: A Longitudinal Study. Inflamm Intest Dis 2021;6(2):109–16. doi: 10.1159/000514576 [published Online First: 20210414]

56. Serrano E, Bastard JP, Trystram L, et al. Serum Versus Fecal Calprotectin Levels in Patients with Severe Obesity Before and 6 Months After Roux-Y-Gastric Bypass: Report of the Prospective Leaky-Gut Study. Obes Surg 2023;33(12):4017–25. doi: 10.1007/s11695-023-06911-w [published Online First: 20231104]

57. Liu T, Zhang L, Joo D, et al. NF-κB signaling in inflammation. Signal Transduct Target Ther 2017;2:17023-. doi: 10.1038/sigtrans.2017.23 [published Online First: 20170714]

58. Atreya I, Atreya R, Neurath MF. NF-kappaB in inflammatory bowel disease. J Intern Med 2008;263(6):591–6. doi: 10.1111/j.1365-2796.2008.01953.x

59. Sokol H, Pigneur B, Watterlot L, et al. Faecalibacterium prausnitzii is an anti-inflammatory commensal bacterium identified by gut microbiota analysis of Crohn disease patients. Proc Natl Acad Sci U S A 2008;105(43):16731–6. doi: 10.1073/pnas.0804812105 [published Online First: 20081020]

60. Rahman MM, McFadden G. Modulation of NF-κB signalling by microbial pathogens. Nat Rev Microbiol 2011;9(4):291–306. doi: 10.1038/nrmicro2539 [published Online First: 20110308]

61. Giri R, Hoedt EC, Khushi S, et al. Secreted NF-κB suppressive microbial metabolites modulate gut inflammation. Cell Rep 2022;39(2):110646. doi: 10.1016/j.celrep.2022.110646

62. Igwe JK, Surapaneni PK, Cruz E, et al. Bariatric Surgery and Inflammatory Bowel Disease: National Trends and Outcomes Associated with Procedural Sleeve Gastrectomy vs Historical Bariatric Surgery Among US Hospitalized Patients 2009-2020. Obes Surg 2023;33(11):3472–86. doi: 10.1007/s11695-023-06833-7 [published Online First: 20231007]

63. Kiasat A, Granström AL, Stenberg E, et al. The risk of inflammatory bowel disease after bariatric surgery. Surg Obes Relat Dis 2022;18(3):343–50. doi: 10.1016/j.soard.2021.12.014 [published Online First: 20211217]

64. Braga Neto MB, Gregory M, Ramos GP, et al. De-novo Inflammatory Bowel Disease After Bariatric Surgery: A Large Case Series. J Crohns Colitis 2018;12(4):452–57. doi: 10.1093/ecco-jcc/jjx177

65. Glassner KL, Abraham BP, Quigley EMM. The microbiome and inflammatory bowel disease. J Allergy Clin Immunol 2020;145(1):16–27. doi: 10.1016/j.jaci.2019.11.003 [published Online First: 2020/01/09]

66. Ramamoorthy S, Levy S, Mohamed M, et al. An ambient-temperature storage and stabilization device performs comparably to flash-frozen collection for stool metabolomics in infants. BMC Microbiol 2021;21(1):59. doi: 10.1186/s12866-021-02104-6 [published Online First: 2021/02/24]

67. FastQC: A Quality Control Tool for High Throughput Sequence Data [Online] [program]. 0.11.9 version: Babraham Bioinformatics, 2010.

68. Ewels P, Magnusson M, Lundin S, et al. MultiQC: summarize analysis results for multiple tools and samples in a single report. Bioinformatics 2016;32(19):3047–8. doi: 10.1093/bioinformatics/btw354 [published Online First: 20160616]

69. Angelova A DD, Subramanian P, Quiñones M, Dolan M, Hurt DE. WGSA2 workflow - a tutorial. protocolsio 2023 doi: doi:10.17504/protocols.io.n92ldm98xl5b/v1

70. Weber N, Liou D, Dommer J, et al. Nephele: a cloud platform for simplified, standardized and reproducible microbiome data analysis. Bioinformatics 2018;34(8):1411–13. doi: 10.1093/bioinformatics/btx617

71. Chen S, Zhou Y, Chen Y, et al. fastp: an ultra-fast all-in-one FASTQ preprocessor. Bioinformatics 2018;34(17):i884–i90. doi: 10.1093/bioinformatics/bty560

72. Wood DE, Lu J, Langmead B. Improved metagenomic analysis with Kraken 2. Genome Biol 2019;20(1):257. doi: 10.1186/s13059-019-1891-0 [published Online First: 20191128]

73. Nurk S, Meleshko D, Korobeynikov A, et al. metaSPAdes: a new versatile metagenomic assembler. Genome Res 2017;27(5):824–34. doi: 10.1101/gr.213959.116 [published Online First: 20170315]

74. Langmead B, Salzberg SL. Fast gapped-read alignment with Bowtie 2. Nat Methods 2012;9(4):357–9. doi: 10.1038/nmeth.1923 [published Online First: 20120304]

75. Cantalapiedra CP, Hernández-Plaza A, Letunic I, et al. eggNOG-mapper v2: Functional Annotation, Orthology Assignments, and Domain Prediction at the Metagenomic Scale. Mol Biol Evol 2021;38(12):5825–29. doi: 10.1093/molbev/msab293

76. Zhu Q, Fisher SA, Shallcross J, et al. VERSE: a versatile and efficient RNA-Seq read counting tool. bioRxiv 2016:053306. doi: 10.1101/053306

77. Zheng J, Ge Q, Yan Y, et al. dbCAN3: automated carbohydrate-active enzyme and substrate annotation. Nucleic Acids Res 2023;51(W1):W115–w21. doi: 10.1093/nar/gkad328

78. Camargo AP, Roux S, Schulz F, et al. Identification of mobile genetic elements with geNomad. Nat Biotechnol 2024;42(8):1303–12. doi: 10.1038/s41587-023-01953-y [published Online First: 20230921]

79. Zhu Q, Fisher S, Shallcross J, et al. VERSE: a versatile and efficient RNA-Seq read counting tool: bioRxiv, 2016.

80. Nayfach S, Camargo AP, Schulz F, et al. CheckV assesses the quality and completeness of metagenome-assembled viral genomes. Nat Biotechnol 2021;39(5):578–85. doi: 10.1038/s41587-020-00774-7 [published Online First: 20201221]

81. Shaffer M, Borton MA, McGivern BB, et al. DRAM for distilling microbial metabolism to automate the curation of microbiome function. Nucleic Acids Res 2020;48(16):8883–900. doi: 10.1093/nar/gkaa621

82. Steinegger M, Söding J. MMseqs2 enables sensitive protein sequence searching for the analysis of massive data sets. Nat Biotechnol 2017;35(11):1026-28. doi: 10.1038/nbt.3988 [published Online First: 20171016]

83. Guo J, Bolduc B, Zayed AA, et al. VirSorter2: a multi-classifier, expert-guided approach to detect diverse DNA and RNA viruses. Microbiome 2021;9(1):37. doi: 10.1186/s40168-020-00990-y [published Online First: 2021/02/02]

84. Roux S, Camargo AP, Coutinho FH, et al. iPHoP: An integrated machine learning framework to maximize host prediction for metagenome-derived viruses of archaea and bacteria. PLoS Biol 2023;21(4):e3002083. doi: 10.1371/journal.pbio.3002083 [published Online First: 20230421]

85. McMurdie PJ, Holmes S. phyloseq: an R package for reproducible interactive analysis and graphics of microbiome census data. PLoS One 2013;8(4):e61217. doi: 10.1371/journal.pone.0061217 [published Online First: 20130422]

86. Kuznetsova A, Brockhoff, P. B., & Christensen, R. H. B. lmerTest Package: Tests in Linear Mixed Effects Models. Journal of Statistical Software 2017;82(13):1–26. doi: 10.18637/jss.v082.i13

87. Oksanen J BF, Kindt R, Legendre P, Minchin P, O’Hara B, Simpson G, Solymos P, Stevens H, Wagner H. Vegan: Community Ecology Package. 2022

88. Wickham H. ggplot2: Elegant Graphics for Data Analysis.: Wickham, H. ggplot2: Elegant Graphics for Data Analysis. 2016.

89. Mallick H, Rahnavard A, McIver LJ, et al. Multivariable association discovery in population-scale meta-omics studies. PLoS Comput Biol 2021;17(11):e1009442. doi: 10.1371/journal.pcbi.1009442 [published Online First: 20211116]

90. Langfelder P, Horvath S. WGCNA: an R package for weighted correlation network analysis. BMC Bioinformatics 2008;9:559. doi: 10.1186/1471-2105-9-559 [published Online First: 20081229]

91. Rahnavard A, Mann B, Giri A, et al. Metabolite, protein, and tissue dysfunction associated with COVID-19 disease severity. Sci Rep 2022;12(1):12204. doi: 10.1038/s41598-022-16396-9 [published Online First: 20220716]

92. Zeng M, Cao H. Fast quantification of short chain fatty acids and ketone bodies by liquid chromatography-tandem mass spectrometry after facile derivatization coupled with liquid-liquid extraction. J Chromatogr B Analyt Technol Biomed Life Sci 2018;1083:137–45. doi: 10.1016/j.jchromb.2018.02.040 [published Online First: 2018/03/17]

93. Jaochico A, Sangaraju D, Shahidi-Latham SK. A rapid derivatization based LC-MS/MS method for quantitation of short chain fatty acids in human plasma and urine. Bioanalysis 2019;11(8):741–53. doi: 10.4155/bio-2018-0241 [published Online First: 2019/04/18]

94. McCloskey D, Gangoiti JA, Palsson BO, et al. A pH and solvent optimized reverse-phase ion-paring-LC–MS/MS method that leverages multiple scan-types for targeted absolute quantification of intracellular metabolites. Metabolomics 2015;11(5):1338–50. doi: 10.1007/s11306-015-0790-y

95. Prinville V, Ohlund L, Sleno L. Targeted Analysis of 46 Bile Acids to Study the Effect of Acetaminophen in Rat by LC-MS/MS. Metabolites 2020;10(1) doi: 10.3390/metabo10010026 [published Online First: 2020/01/16]

96. Sergushichev AA. An algorithm for fast preranked gene set enrichment analysis using cumulative statistic calculation. bioRxiv 2016:060012. doi: 10.1101/060012

97. Cheung F, Fantoni G, Conner M, et al. Web Tool for Navigating and Plotting SomaLogic ADAT Files. J Open Res Softw 2017;5 doi: 10.5334/jors.166 [published Online First: 20170908]

98. Kozich JJ, Westcott SL, Baxter NT, et al. Development of a dual-index sequencing strategy and curation pipeline for analyzing amplicon sequence data on the MiSeq Illumina sequencing platform. Appl Environ Microbiol 2013;79(17):5112–20. doi: 10.1128/aem.01043-13 [published Online First: 20130621]

99. Callahan BJ, McMurdie PJ, Rosen MJ, et al. DADA2: High-resolution sample inference from Illumina amplicon data. Nat Methods 2016;13(7):581–3. doi: 10.1038/nmeth.3869 [published Online First: 2016/05/24]

100. Rohart F, Gautier B, Singh A, et al. mixOmics: An R package for ‘omics feature selection and multiple data integration. PLoS Comput Biol 2017;13(11):e1005752. doi: 10.1371/journal.pcbi.1005752 [published Online First: 20171103]

