## Supplementary figures and images for "Gut microbiome shifts in adolescents after sleeve gastrectomy with increased oral-associated taxa and pro-inflammatory potential"

### Supplemental Figure 1

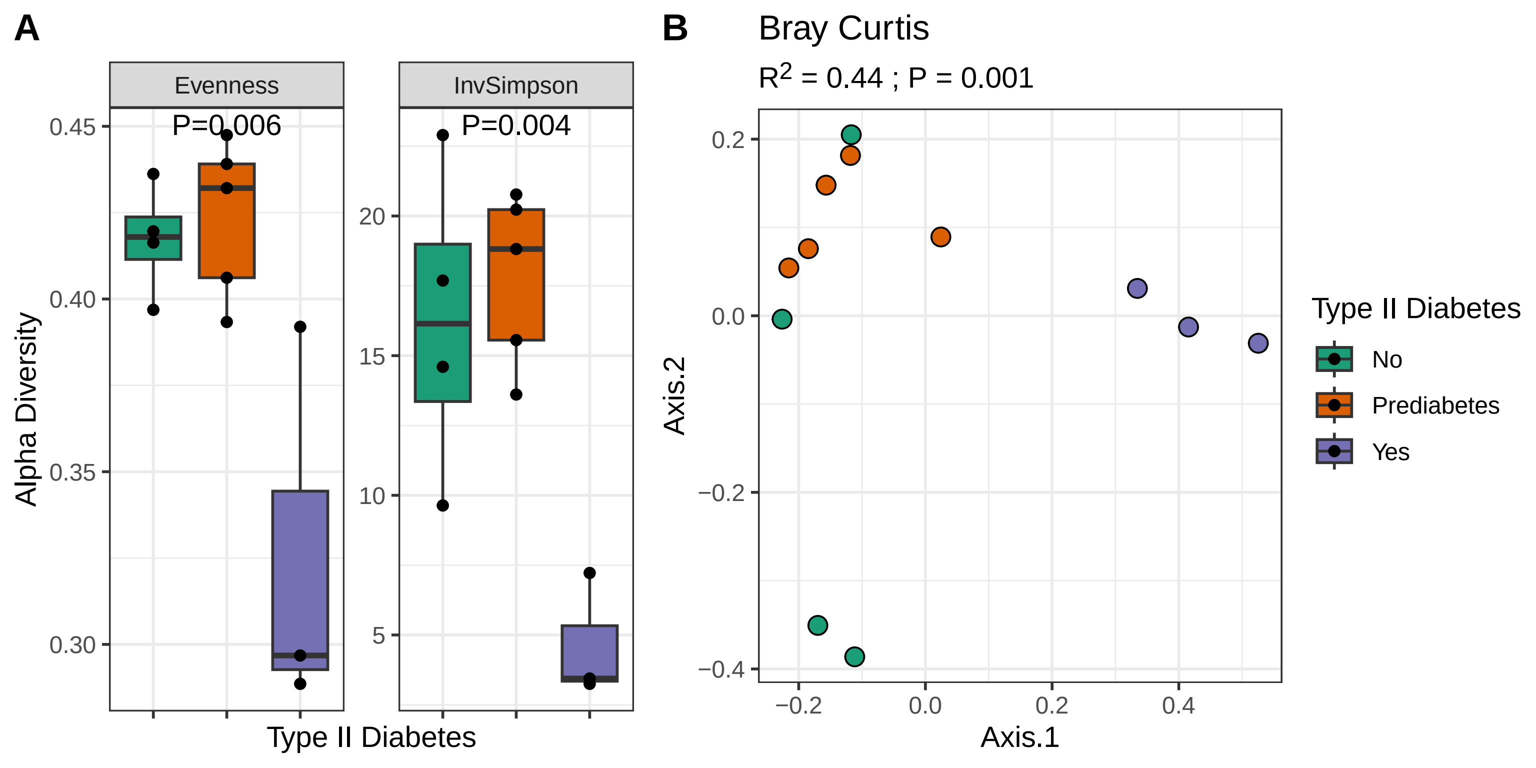

### Supplemental Figure 2

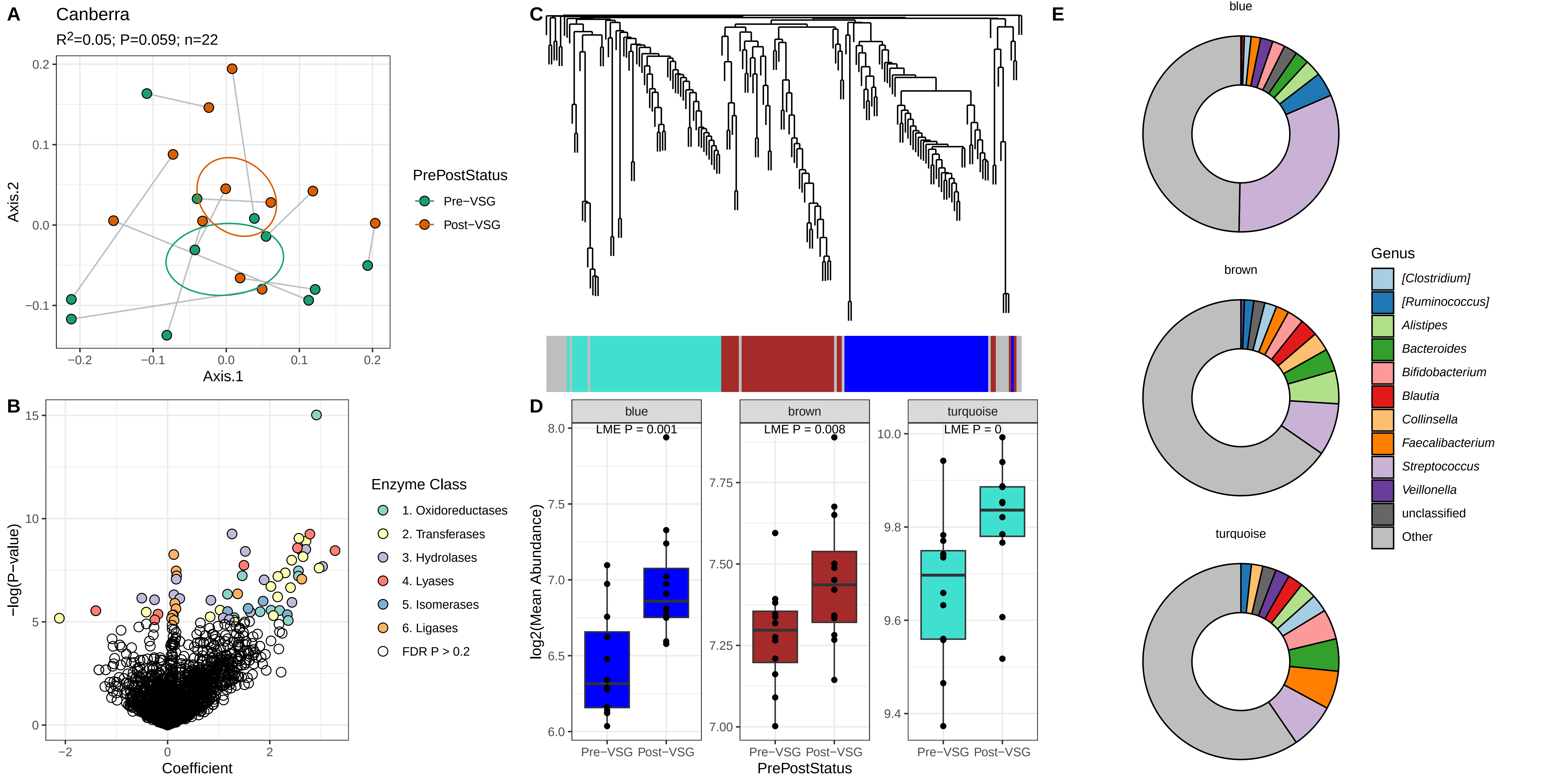

### Supplemental Figure 3

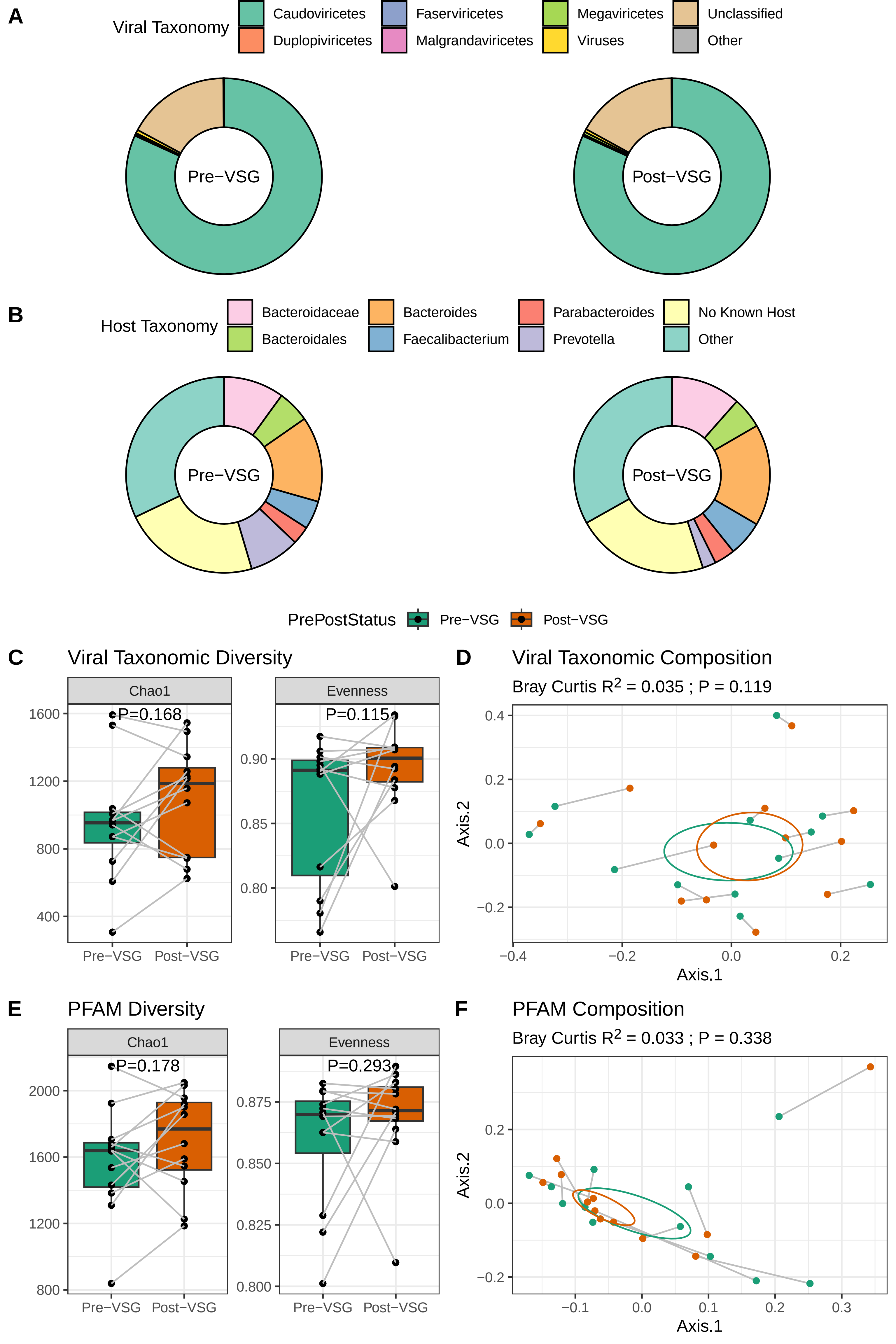

### Supplemental Figure 4

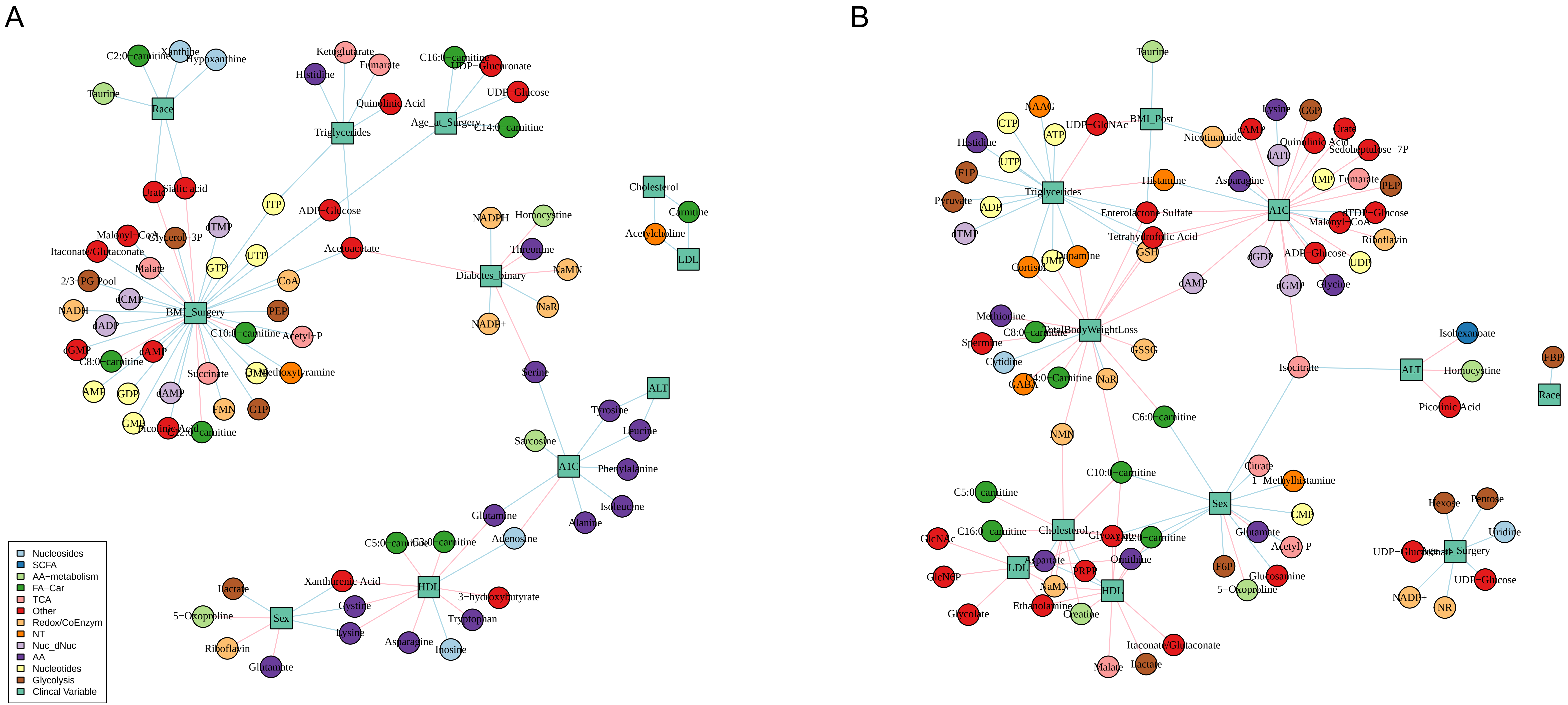

### Supplemental Figure 5

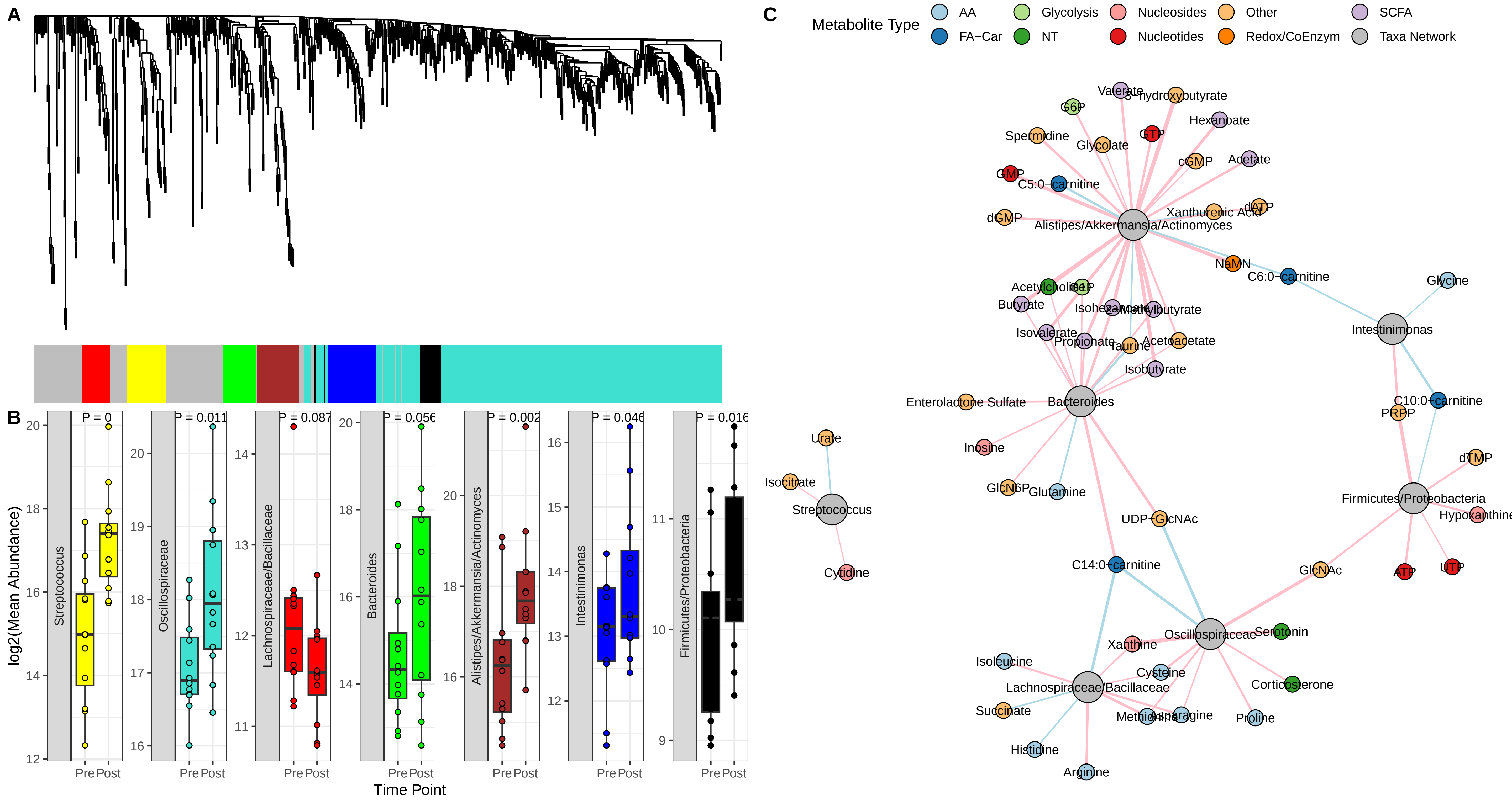

### Supplemental Figure 6

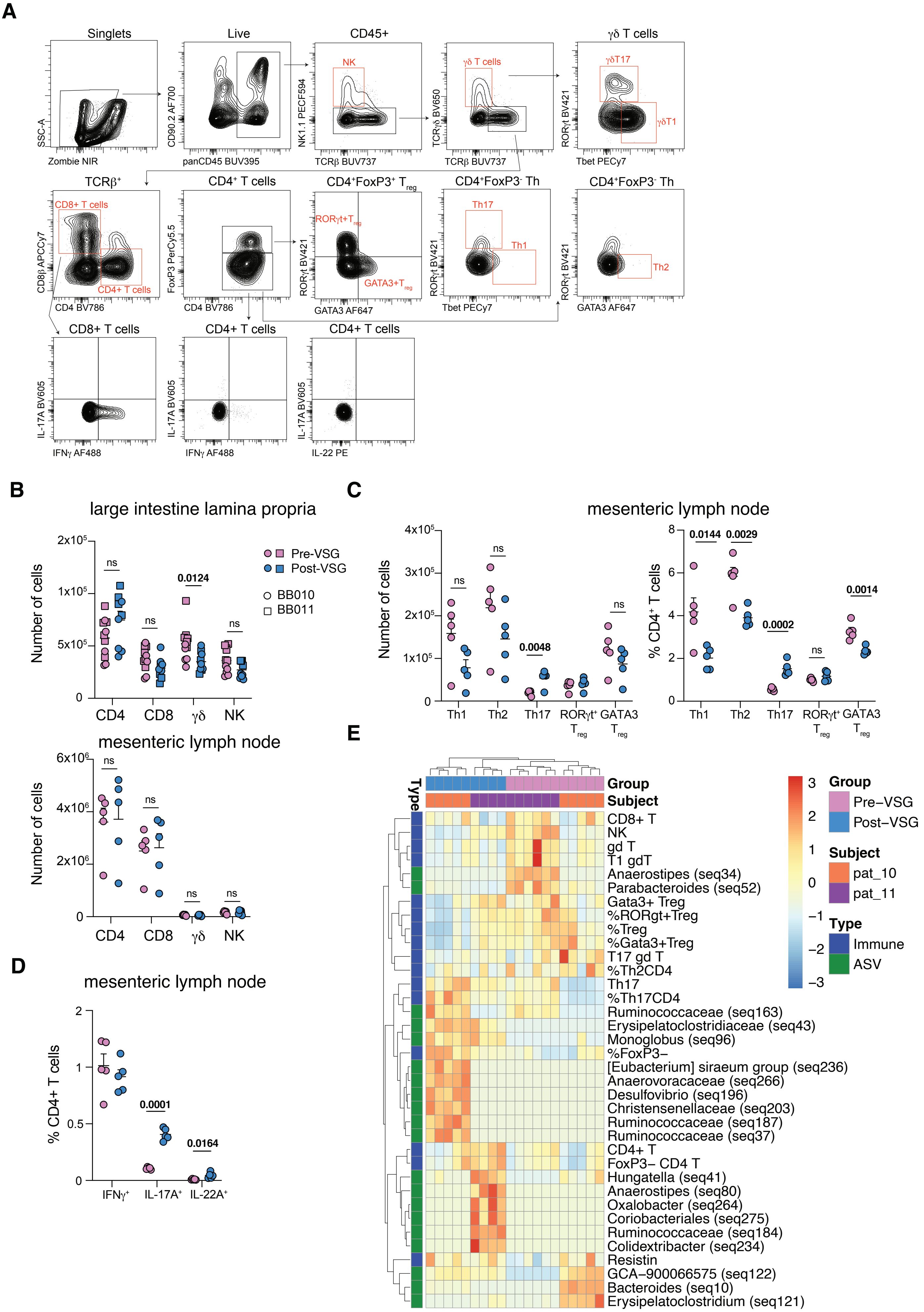

### Supplemental Figure 7

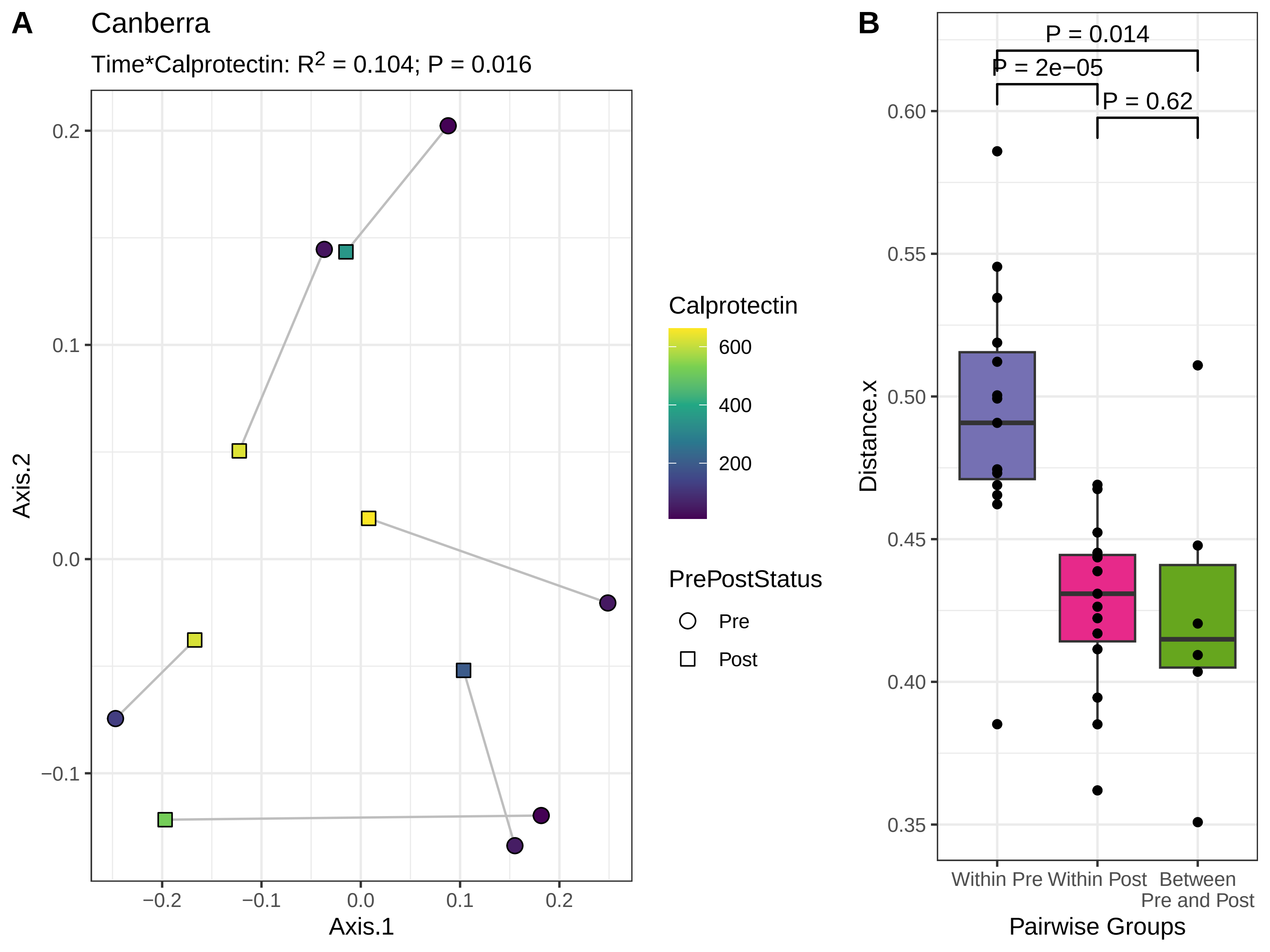

### Supplemental Figure 8

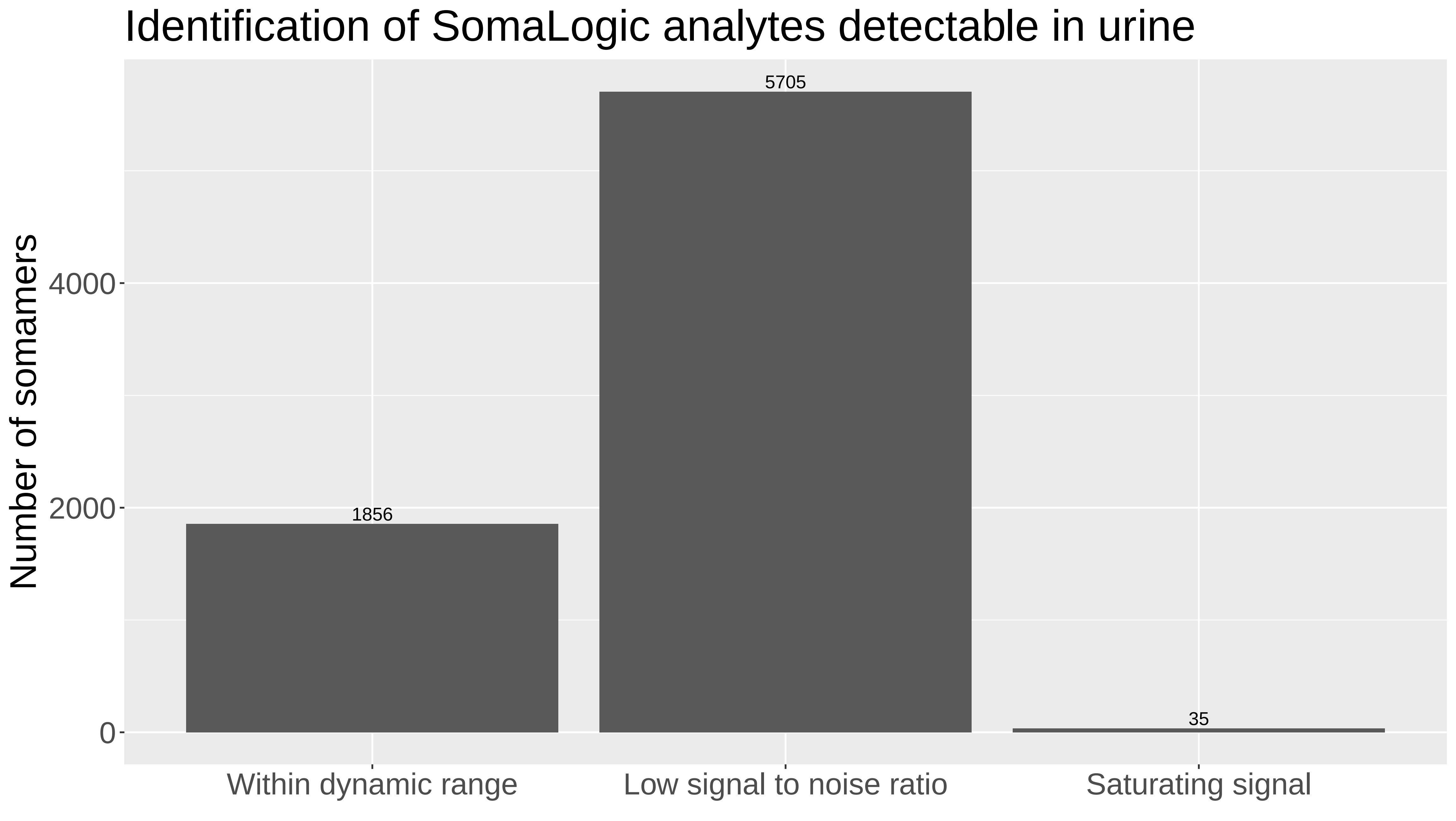
